# Comparison of gait speed reserve, usual gait speed, and maximum gait speed of adults aged 50+ in Ireland using explainable machine learning

**DOI:** 10.1101/2021.07.23.21260911

**Authors:** James R.C. Davis, Silvin P. Knight, Orna A. Donoghue, Belinda Hernández, Rose Anne Kenny, Roman Romero-Ortuno

## Abstract

Gait speed is a measure of general fitness. Changing from usual (UGS) to maximum (MGS) gait speed requires a general effort across many body systems. The difference, MGS – UGS, is defined as gait speed reserve (GSR). In the present study, using 3925 participants aged 50+ from Wave 3 of The Irish Longitudinal Study on Ageing (TILDA), we used a gradient boosted trees-based stepwise feature selection pipeline for the discovery of clinically relevant predictors of GSR, UGS, and MGS using a shortlist of 88 features across 5 categories (socio-demographics/anthropometrics/medical history; cardiovascular system; physical strength; sensory; and cognitive/psychological). The TreeSHAP explainable machine learning package was used to analyse the input-output relationships of the three models.

The mean 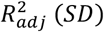 from 5-fold cross validation on training data and the 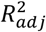 score on test data for the models are: 0.38 (0.04) and 0.41 for UGS; 0.45 (0.04) and 0.46 for MGS; and 0.19 (0.02) and 0.21 for GSR.

Features selected for the UGS model were: age, chair stands time, body mass index, grip strength, number of medications, resting state pulse interval, mean motor reaction time in the choice reaction time test, height, depression score, sit-to-stand difference in diastolic blood pressure, and left visual acuity.

The features selected for the MGS model were: age, grip strength, repeated chair stands time, body mass index, education, mean motor reaction time in the choice reaction time test, number of medications, height, the standard deviation of the mean reaction time in the sustained attention to response task, mean heart rate at resting state, fear of falling, MOCA errors, orthostatic intolerance during active stand, smoking status, total heart beat power during paced breathing, the root mean square of successive differences between heartbeats during paced breathing, and visual acuity.

Finally, the features chosen for the GSR model were: mean motor reaction time in the choice reaction time test, grip strength, education, chair stands time, MOCA errors, accuracy proportion in the sound induced flash illusion (two beeps and one flash with stimulus-onset asynchrony of +150 ms), fear of falling, height, age, sex, orthostatic intolerance, MMSE errors, and number of cardiovascular conditions.

MGS and UGS were more explainable than GSR. All three models contain features from all five categories. There were common features to all three models (age, grip strength, chair stands time, mean motor reaction time in the choice reaction time test, and height), but also some features unique to each of them. Overall, findings on all three models were clinically plausible and support a network physiology approach to the understanding of predictors of performance-based tasks. By employing an explainable machine learning technique, our observations may help clinicians gain new insights into the multisystem predictors of gait speed and gait speed reserve in older adults.

## 1 Introduction

Gait speed is a measure of general fitness (1); faster gait speed is associated with the ability to meet occupational demands in younger adults (2), while slower gait speed is associated with functional decline and morbidity in older adults (3, 4). Even though usual (or comfortable) walking speed and maximum walking speed are significantly intercorrelated (5), changing from comfortable to maximum speed requires a general effort across many body systems. The difference between these two gait speeds has been referred to as gait speed reserve (6).

Gait speed reserve (GSR) may be a useful proxy measure of physiological reserve in humans. For example, some studies have suggested that in community-dwelling older adults, the simultaneous consideration of both usual and maximum gait speed could increase the accuracy of the identification of frailty (7, 8). Frailty is generally understood as a state of reduced physiological reserve, increasing the risk of decompensation and adverse clinical outcomes triggered by relatively minor stressors (9, 10); however, there is still no universal agreement as to how to operationalize frailty in clinical practice and research (11). Consequently, there is increasing scientific and medical interest to understand what the main determinants of physiological reserve are during specific performance-based tasks, and the study of objective ways in which to quantify and predict that reserve (12, 13). Although this is a grand challenge, research has suggested that a way to deepen our understanding of physiological reserve in older adults is through the measurement of organ function across many systems (14).

To our knowledge, there have been no systematic attempts to model predictors of GSR in a large representative sample of community-dwelling older adults where many demographic, anthropometric and clinical features are measured across multiple physiological systems.

In this study, we aimed to use machine learning to: first, identify the set of features that, from a shortlist of features, best describe UGS, MGS, and GSR; then, using explainable machine learning methods, look into the selected models for UGS, MGS, and GSR to observe how each feature in the model was associated with the output in a non-parametric manner. With the selected features and visualizations of the input-output relationships we then discussed the clinical interpretations with respect to the cohort used and the hypothesis that UGS, MGS and GSR are multisystem phenomena.

In this paper, we first describe the TILDA study and the methods used to collect data on the shortlisted features. Next, we describe the feature selection pipeline, and provide an overview of the histogram gradient boosting regression machine learning model employed. We end the materials and methods section with a brief description of the Shapley additive explanations (SHAP) package used to explain the models. The results from feature selection, and SHAP interpretation are then presented separately for each of the three models: UGS, MGS, and GSR. Finally, the discussion and conclusions compare the results of each model, and comment on the potential clinical relevance.

## 2 Materials and Methods

### 2.1 Design and Setting

We analyzed data from adults aged 50+ from Wave 3 of TILDA, a population-based longitudinal study of ageing (https://tilda.tcd.ie/). TILDA study design, as well as the full cohort profile, have been previously described in detail (16, 17). Wave 3 data collection took place in 2014 and 2015 and included a computer-assisted personal interview conducted by social interviewers in the participants’ home, a self-completion questionnaire completed in the participants’ own time and a detailed suite of technology-aided health assessments conducted by trained research nurses at a dedicated health assessment centre. Ethical approval was obtained from the Faculty of Health Sciences Research Ethics Committee at Trinity College Dublin, Ireland (Reference: Main Wave 3 Tilda Study; approval date: 9^th^ June 2014). All participants provided written informed consent and all data collection procedures adhered to the World Medical Association Declaration of Helsinki on ethical principles for medical research involving human subjects.

#### 2.1.1 Analytical Sample

The primary analytical sample consisted of participants from TILDA Wave 3 aged 50 years or more who had data for both UGS and MGS.

#### 2.1.2 Gait Speed Measures

At Wave 3 of TILDA, gait speed was measured as part of a health centre assessment. Measurements in units of cm/s were made using a 4.88 m computerized walkway (GAITRite, CIR Systems, NY, USA). A two-meter space before and after the walkway was used for acceleration and deceleration. Participants were first asked to walk at their normal (usual) pace (usual gait speed: UGS), and then as fast as they safely could (maximum gait speed: MGS). Two walking trials were obtained in each condition and the mean value for each was used in this analysis. GSR was defined as MGS – UGS.

#### 2.1.3 Shortlisted Features

A shortlist of features from the TILDA Wave 3 dataset was manually curated by the lead author (JD, trained in a STEM discipline), in consultation with co-authors representing both STEM (RR, BH) and health/medical (SK, OD, RRO) fields. The features were chosen based on known or plausible associations with the three gait speed modalities under investigation. The feature curation considered features from the following five categories: socio-demographics/anthropometrics/medical history, cardiovascular system, physical strength, senses, and cognitive/psychological.

##### 2.1.3.1 Socio-demographics/Anthropometrics/Medical History

Demographic information included **Age** in years, **Sex** (male=0; female=1), and level of educational attainment (**Edu3**): either primary/none (Edu3=1), secondary (Edu3=2), or tertiary/higher (Edu3=3).

Anthropometrics comprised **Weight** (cm), **Height** (cm), body mass index (**BMI**, kg/m^2^), and waist-to-hip ratio (**WaistHipRatio**: waist circumference / hip circumference) (18).

Medical history: number of cardiovascular diseases (**nCVD**, from the following list: hypertension; angina; heart attack; congestive heart failure; diabetes; stroke; transient ischaemic attack; high cholesterol; heart murmur; abnormal heart rhythm; atrial fibrillation), if taking any **Antidepressant** medications (binary), if taking any **Antihypertensive** medications (binary), and the total number of medications being taken excluding supplements (**nMeds**) (19).

Smoking status (**Smoker**) was categorized as: never (smoker=0); past (smoker=1); and current (smoker=2). Alcohol intake was scored with the **CAGE** scale (20).

The number of reported difficulties with activities of daily living were also assessed. The six basic activities (**ADLs**) were: dressing, including putting on shoes and socks; walking across a room; bathing or showering; eating, such as cutting up food; getting in or out of bed; and using the toilet, including getting up or down. The six independent activities (**IADLs**) were: preparing a hot meal; doing household chores (laundry, cleaning); shopping for groceries; making telephone calls; taking medications; and managing money such as paying bills and keeping track of expenses (21).

##### 2.1.3.2 Cardiovascular System

During the TILDA Wave 3 health assessment resting state (RS) cardiovascular measurements were made during an approximately 10-minute window in which the participant was laying supine in a comfortably lit room at an ambient temperature of between 21 °C and 23 °C. The full TILDA active stand protocol in which the resting state window takes place has been detailed elsewhere (22-24). Throughout the RS participants underwent non-invasive continuous haemodynamic monitoring using a Finometer MIDI device (Finapres Medical Systems BV, Amsterdam, the Netherlands). All RS parameters selected for the shortlist are mean values from the last minute of supine rest (22). Hemodynamic parameters were: systolic blood pressure (**sBP_RS**), diastolic blood pressure (**dBP_RS**), mean arterial pressure (**MAP_RS**) all in units of mmHg, heart rate (**HR_RS**) in bpm, stroke volume (**StrokeVolume_RS**) in mL, left ventricular ejection time (**LVET_RS**) in ms, pulse interval (**PulseInterval_RS**) in ms, maximum slope (**Maxslope_RS**) in mmHg/s, cardiac output (**CardiacOutput_RS**) in L/min, and total peripheral resistance (**TPR_RS**) in dyn ⋅ s ⋅ cm^−5^. A near-infrared spectroscopy (NIRS) device, attached over the participants’ left frontal lobe area, was also employed during the RS and the following cerebral oxygenation features were extracted, again as the mean values from the final minute of rest: oxygenated haemoglobin concentration (**O2Hb_RS**) and deoxygenated haemoglobin concentration (**HHb_RS**) both in units of μMol/L, and tissue saturation index (**TSI_RS**) as a percentage (22). Previously derived sample entropy values for resting sBP (**sBP_RS_SampEn**), dBP (**dBP_RS_SampEn**), MAP (**MAP_RS_SampEn**), heart rate (**HR_RS_SampEn**), O2Hb (**O2Hb_RS_SampEn**), HHb (**HHb_RS_SampEn**), and TSI (**TSI_RS_SampEn**) were also shortlisted (22). In addition, participants were asked if they experienced dizziness upon standing (**PhasicDizziness**: yes or no), and this feature was also included in the shortlist.

Resting heart rate variability measures were also shortlisted; these were obtained in two five-minute blocks as detailed elsewhere (25). In short, for each block, participants were laying supine. In the first block, participants were asked to breath spontaneously (free breathing), and in the second block, they were asked to breathe according to a pre-recorded set of auditory instructions (paced breathing at a frequency of 0.2 Hz). Measurements were obtained using 3-lead electrocardiograms (Medilog Darwin, Oxford Instruments Medical Ltd, UK). The data were subject to a 0.01 – 1000 Hz band-pass filtering before R peak detection was performed with a proprietary software (26). The data collection and processing are described in detail elsewhere (25). Time domain features were: mean heart rate in bpm, root-mean-square of successive differences between RR intervals in ms, standard deviation of NN intervals in ms, and difference between maximum and minimum heart rate in bpm, derived for both free (**HR_Mean_Free, HR_rMSSD_Free, HR_SDNN_Free, HR_Span_Free**) and paced breathing (**HR_Mean_Paced, HR_rMSSD_Paced, HR_SDNN_Paced, HR_Span_Paced**). The difference between free and paced breathing values was calculated for rMSSD (**HR_rMSSD_PacedFreeDiff**). In the frequency domain total spectral power in the 0 - 0.4 Hz frequency band was measured for both free (**HR_TotalPower_Free**) and paced breathing (**HR_TotalPower_Paced**) in units of milliseconds squared, ms^2^.

sBP, dBP and HR were also determined in a more conventional manner using a sphygmomanometer in seated (**sBP_Seated, dBP_Seated**, and **HR_Seated**) and standing (**sBP_Standing, dBP_Standing**, and **HR_Standing**) positions; all with units of mmHg. The difference between seated and standing values were calculated for each of the measures (**sBP_SeatStandDiff, dBP_SeatStandDiff**, and **HR_SeatStandDiff**).

Pulse wave velocity (**PulseWaveVelocity**), a non-invasive measure of arterial stiffness with units of m/s, was also included as a cardiovascular feature. In TILDA, the average of two measurements between the carotid and femoral arteries (in m/s) was obtained using a Vicorder^®^ (SMT medical GmbH & Co. Wuerzburg, Germany). Full details have been described elsewhere (16, 18).

##### 2.1.3.3 Physical Strength

Upper and lower body strength were assessed via grip strength and chair stands time. Grip strength was measured in kg using a hydraulic hand dynamometer (Baseline^®^, Fabrication Enterprises, Inc., White Plains, NY, USA). The value for grip strength referred to henceforth (**GripStrength**) is the maximum value from a total of eight measurements with four made on each hand. Lower body strength was assessed using the chair stands test in which the time (in seconds) was recorded for the participants to complete five chair stands as quickly as possible, keeping the arms folded across their chest (**ChairStandsTime**). Chair height was 46 cm.

##### 2.1.3.4 Cognitive and Psychological

Global cognition was assessed using two paper-based assessments: the Montreal Cognitive Assessment (MOCA) (27) and the Mini-Mental State Examination (MMSE) (28); from these, the number of errors (**MOCA_errors, MMSE_errors**) were extracted for the feature shortlist. Concentration, cognitive processing, and motor response were assessed using two computer assisted tasks: the choice reaction task (29) and the sustained attention to response task (SART) (30). The choice reaction task required participants to hold down a central button until an on-screen stimulus (either the word “YES” or “NO”) appeared, at which time they had to press the corresponding button on a keyboard. After pressing either button, participants were then required to return to the central button to continue. This was repeated approximately 100 times. In the SART test, participants watched a screen that displayed the numbers 1 – 9 sequentially a total of 23 times. A number appeared for 300 ms with an interval of 800 ms between numbers: the entire trial lasts approximately four minutes. Participants were instructed to keep a central button pressed until observing a specific number (i.e. 3), at which point they were required to press a different button and then return to the central button. We extracted the following features from the choice reaction task: mean and standard deviation of cognitive reaction time (**CRT_mean, CRT_SD**) and motor response time (**MRT_mean, MRT_SD**), and the number of correct CRT presses (**CRT_correct**). CRT is the time taken to release the central button in response to the stimulus; MRT is the time between releasing the central button and pressing the required button. From the SART, we extracted: mean and standard deviation of reaction time (**SART_mean, SART_SD**), and number of trails in which the participant pressed when the number 3 appeared (**SART_errors**). CRT, MRT, and SART times are all measured in milliseconds.

The psychological domains of depression, anxiety, and loneliness were assessed using the Center for Epidemiologic Studies Depression Scale (**CESD**), the Hospital Anxiety and Depression Scale – Anxiety subscale (**HADSA**), and the UCLA Loneliness Scale (**UCLA**), respectively. Fear of falling (**FOF**) was determined with a yes or no question (16).

##### 2.1.3.5 Sensory

Visual acuity (VA) was measured using a LogMar chart. VA in the left eye (**VisualAcuityLeft**), right eye (**VisualAcuityRight**), and best VA (**VisualAcuityBest**) were included in this work. VA left and right were in logarithmic units. Best VA was defined as 100 − (min ([*VA*_*left*_, *VA*_*right*_]) × 50. Contrast sensitivity (CS) was measured at five spatial frequencies; in cycles per degree (cpd) they were: 1.5 cpd (**cs_score_a**), 3 cpd (**cs_score_b**), 6 cpd (**cs_score_c**), 12 cpd (**cs_score_d**), and 18 cpd (**cs_score_e**). The procedures for visual acuity and contrast sensitivity measurements are described in detail elsewhere (31). Self-reported hearing (**Hearing_SR**) was ascertained by the question: “Is your hearing (with or without a hearing aid): 1. Excellent, 2. Very good, 3. Good, 4. Fair, or, 5. Poor?”

Multisensory integration was measured using the Shams sound-induced flash illusion (SIFI) test (32). The procedure used in TILDA is described in more detail elsewhere (33); but in short, participants were subjected to a set of beeps and flashes and asked to report how many flashes they perceived. Five general flash-beep combinations were presented to the participants: 2 beeps + two flashes; 1 beep + 1 flash; 0 beeps + 1 flash; 0 beeps + 2 flashes; and 2 beeps + 1 flash. The flash-beep configurations used in this analysis are the so-called ‘illusory’ 2 beep 1 flash (2B1F) trials. In 2B1F trials, the flash is synchronous with one of the beeps; the other beep occurred either 70 ms, 150 ms, or 230 ms before (**SIFI_2B1F_70, SIFI_2B1F_150, SIFI_2B1F_230**) or after (**SIFI_2B1F_m70, SIFI_2B1F_m150, SIFI_2B1F_m230**) the flash-beep pair. SIFI susceptibility represented accuracy for judging how many flashes were presented when one flash was presented with two beeps (2B1F). Lower accuracy, judging one flash as two, thus indicates higher SIFI susceptibility and stronger integration. SIFI susceptibility was expressed as proportion correct. As there were two trials per condition, these variables were considered discrete (i.e., participants scored 0, .5 or 1 proportion correct) (34).

#### 2.1.4 Feature Selection Algorithm

All operations were performed using Python 3. The feature selection was executed on the Tinney High Performance Computing Cluster at Trinity College Dublin (https://www.tchpc.tcd.ie/node/1353).

Prior to any feature selection or training of any kind, the data were divided according to an 80/20 train/test split. From the shortlisted set of 88 features across 5 domains, features were chosen for the final models using an automated stepwise feature selection algorithm. In this algorithm, each feature is individually added to a temporary model that contains all features previously selected for the final model: for the initial round, each temporary model contains a single feature. For each individual temporary model, a hyperparameter tuning is performed in which a 5-fold cross validation (CV) is performed on the training data for each set of hyperparameters. The hyperparameter tuning is in the form of a 100-iteration randomized search of a set of predefined hyperparameter distributions.

The evaluation metric employed was the adjusted-*R*^2^metric 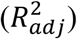. This metric is used to avoid the continual increase in *R*^2^ that occurs with the addition of new features regardless of whether they significantly increase the variance explained by the model. For each temporary model, the best parameters are chosen based on the mean 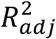 from 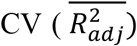. From these temporary models, the one that provides the biggest increase in 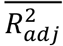 is chosen to continue on with, i.e. the new feature upon which that temporary model is based is added to the final model. Before moving to the selection of the next feature each feature in the model is removed one by one to check if any of them have become redundant in light of the addition of the newest feature; if the score improves on removing a feature then that feature is removed from the model.

For the purpose of performance monitoring, on each iteration of the loop, the current best model is fit to the entire training dataset and evaluated on both the training and test sets to give training and test 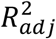 scores. These scores do not influence feature selection.

#### 2.1.5 Histogram Gradient Boosting Regression

The regression model employed for this analysis was the histogram gradient boosting regressor (HGBR) from Sci-kit learn (35). The Sci-kit learn implementation is based on Microsoft’s light gradient boosting machines (36). Gradient boosting (37) is a machine learning technique that builds decision trees sequentially where each one is constructed such that it predicts the residuals from the previous tree. Gradient boosting is a powerful tool that has become the model of choice in many fields and applications (38) and have been shown to outperform deep-learning models where the data are tabular and the features themselves have individual meanings as opposed to data structured in a temporal and/or spatial manner as is the case for problems in image and audio domains (39). Light gradient boosting machines and histogram gradient boosting is an adaptation of gradient boosted trees that places feature values into histogram-like bins which allow for tree split points to be located more efficiently.

The HGBR model inherently supports missing values and categorical data. The support for missing data helps to avoid the need for data imputation or removal of features. The categorical data support avoids the need for dummy variables and one-hot encoding which can drastically increase the dimensionality of the input feature space.

#### 2.1.6 SHAP Values

Two important characteristics of an input feature in a model are: the relationship between the feature and the model output; and the importance of the feature in the model i.e. how much does the feature impact the model. The latter characteristic can be broken down into global impacts: how does the feature effect the model as a whole; and local impacts: how a feature impacts the output of the model for an individual sample.

When using linear models, the feature coefficients readily allow for interpretation of the input-output relationship. Such models also allow for significance statistics to be easily calculated. However, it is also the case that whilst linear models are easy to use and explain, they are very often not able to capture the underlying structure of the data. It has also been shown that linear models can complicate and reduce interpretation by attributing importance to features that in reality have no relationship to the output when the underlying data structure is non-linear (39). Non-linear machine learning models can allow for more complex associations and interactions to be captured but result in a model that is generally harder to interpret. Tree based models are, in theory, very explainable, but when ensembles of trees are present, the interpretability becomes impractical. Methods for the global assessment of features exist (e.g. feature permutation and tree-based impurity) but these approaches judge how much the model as a whole depend on a particular feature and miss out on what impact a certain feature has on a specific sample.

Individual conditional expectation and partial dependence plots have been developed to investigate the input-output relationships, but they are expensive to compute and are thus not widely used. Local Interpretable Model-agnostic Explanations (LIME) (40) were proposed in a paper entitled “Why Should I Trust You?” as a method to investigate local interpretations of any machine learning model. Whilst being an extremely important development in explainable machine, they rely on linear models to make the local interpretations, which may not always be suitable. This may especially be the case when studying human physiology, as its processes are often regulated by nonlinear dynamics (41).

In view of the above considerations, in this work SHapley Additive exPlanations (SHAP) values (42) were employed to assess feature importance and investigate the impact of features on the model output. The TreeExplainer SHAP method builds interpretations that are theoretically guaranteed to be faithful at the local and global levels (39).

Shapley values, from which the SHAP package derives, were presented in the field of cooperative game theory (43). They guarantee a fair distribution of contributions from each feature in a model. However, it is generally NP-hard (i.e. complexity scales exponentially with number of features) to compute them and as such, they have not been widely utilized. The SHAP package first developed a model agnostic heuristic that allowed for their use. A more recent development allows for exact Shapley values to be computed for tree-based models in low-order polynomial time (as opposed to exponential time for the NP-hard problem). In other words, the TreeExplainer package allows for exact Shapley values to be calculated for tree-based models in practical time even for large data.

Detailed derivations of Shapley values and of the TreeExplainer algorithm (39) can be found elsewhere, but a brief and simplified description of Shapley values shall be given here.

The contribution *δ*, of a single feature *i* (from the set of features, *S*), for a sample instance *x*, to a given model, is computed as,

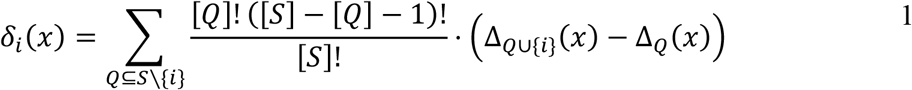

where *Q* is a subset of *S*, and Δ is the contribution of worth to the model from a subset of features.

Shapley derived these values as a method of attributing worth to each player in a game in a fair way. In coalitional game theory *n* players form a grand coalition, S, that has a total worth, Δ_*S*_. Each player is representative of an input feature. Each smaller coalition, *Q* ; *Q* ⊂ *S*, has worth Δ_*Q*_. A Shapley value is a unique solution that satisfies the following four axioms developed to ensure a fair distribution of worth:

1. 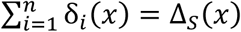 The sum of contributions from each player equals the total worth of the game.
2. ∀*W* ⊆ *S* \ {*i*}: Δ_*W*_ = Δ_*W*∪{*i*}_ ⇒ δ_*i*_(*x*) = 0 If a coalition *W* not containing player *i* has a worth equal to that of coalition *W* in union with player *i*, then the worth of player *i* is zero; i.e. the player *i* did not increase the worth of coalition *W*.
3. ∀*W* ⊆ *S* \ {*i, j*}: Δ_*W*∪{*i*}_ = Δ_*W*∪{*j*}_ ⇒ *δ*_*i*_(*x*) = *δ*_*j*_(*x*) If the worth of *W* in union with player *i* is equal to the worth of *W* in union with *j*, the worth of player *j* is equal to the worth of player *j*.
4. ∀*w, x* ∈ X: *δ*(*w* + *x*) = *δ*(*w*) + *δ*(*x*), *where* Δ_*Q*_(*w* + *x*) = Δ_*Q*_(*w*) + Δ_Q_(*x*), ∀*Q* ⊆ *S* For all instances *w* and *x*, the contribution of a single feature for the sum of values from instances *w* and *x* is equal to the sum of the contributions from that feature having values of *w* and *x*, where the same is true for any subset Q of features and instances w and x.

TreeExplainer is designed such that it does not need to compute Shapley values for the entire feature set but instead uses the tree structure to perform the exact computation on smaller feature sets made possible by the tree.

SHAP values are computed for each instance of each feature. This allows for global feature explanations to be constructed either visually in the form of SHAP summary plots or as a single value such as mean absolute SHAP value, or maximum absolute SHAP value. The nature of SHAP values being true to local impacts of features means that low frequency, high impact effects do not go unnoticed. SHAP interaction values are also readily available that explain the impact of interactions between two features. SHAP values are presented as having a positive or negative impact on the output of the model with respect to the expected model output i.e. the mean output of the model. So, for an individual sample, the SHAP value for a particular feature might be for example, -2.5; this should be interpreted as: the value of that feature for that sample is associated with a model output that is -2.5 units less than the model’s mean output.

## 3 Results

Note on presentation of results: the method for feature selection describes a situation whereby features can be removed from the model if they are made redundant by the addition of new features; this did not occur in any of the models and as such all features named henceforth with regard to feature selection are to be understood as features added to the model.

### 3.1 Analytical Cohort

In TILDA Wave 3, 4309 participants completed the health centre assessment (22), where the gait speed tests were conducted. After exclusion of participants aged less than 50 years or with missing data for either UGS or MGS, there were N = 3925 participants, with 2156 (55%) being female. An analytical sample inclusion flowchart can be seen in Figure 1. The educational attainment breakdown was as follows: third/higher: 1685 (43%), secondary: 1571 (40%), and primary/none: 669 (17%). The analytical cohort had a mean (SD) age of 64.5 (7.8) years, UGS of 136.7 (19.2) cm/s, MGS of 171.0 (26.9) cm/s, and GSR of 34.3 (16.6) cm/s.

**Figure 1:**
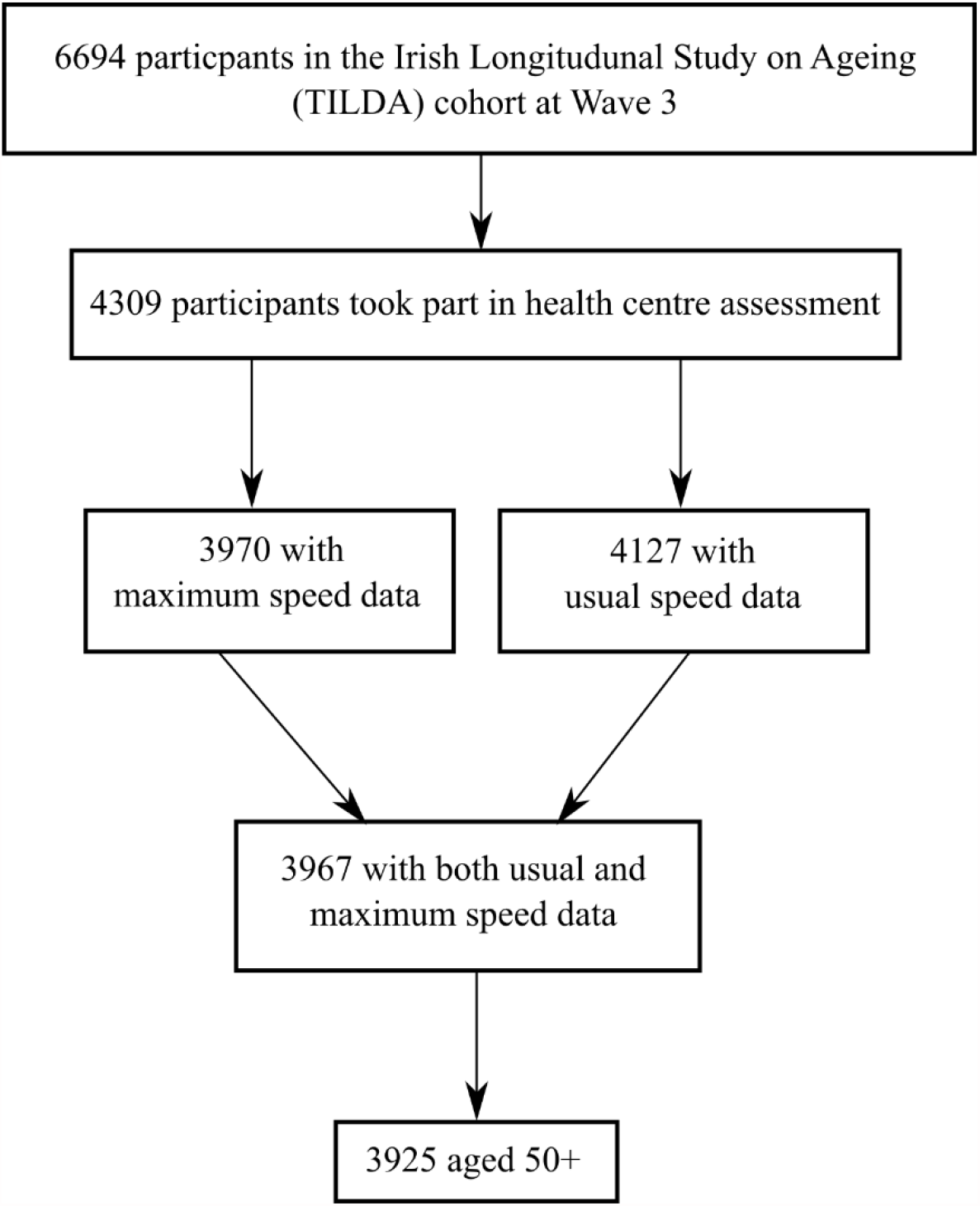
analytical sample inclusion flowchart.

### 3.2 Usual Gait Speed

The peak 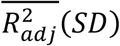 achieved for the UGS model was 0.38 (0.04) with training and test scores of 0.43 and 0.41, respectively. The expected model output was 136.6 cm/s. The features chosen for the model, in order of selection as per Figure 2 were: age, chair stands time, BMI, grip strength, number of medications, resting state pulse interval, mean motor reaction time, height, depression score, sit-to-stand difference in diastolic blood pressure, and left visual acuity.

**Figure 2.**
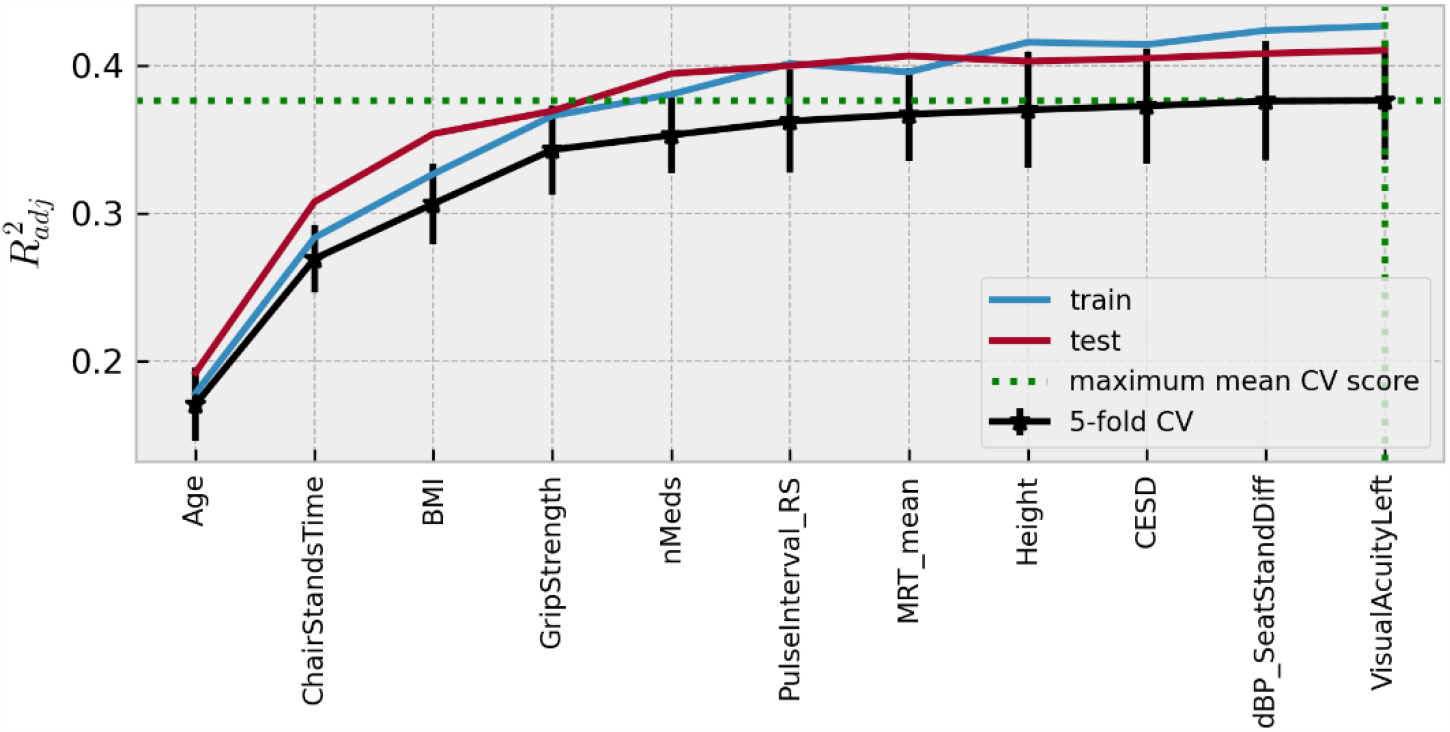
**Visualization of the feature selection process for the usual gait speed model. From left to right on the x-axis, the features are in order of addition to the model. The y-axis shows the dimensionless** 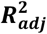 **metric. Mean 5-fold cross-validation scores with error bars showing** ± **SD are shown in black, train scores in blue, and test scores in red. The feature at which the peak score was achieved is highlighted by the green dotted lines.**

A SHAP summary plot is shown in Figure 3; each point on the x-coordinate represents a samples SHAP value, and its colour signifies the value of the feature for that sample, with red being high, blue being low and nan (missing) values appearing grey. On the y-axis, features are arranged from top to bottom in order of decreasing mean absolute SHAP value: chair stands time, age, body mass index, number of medications, grip strength, resting state pulse interval, height, mean motor reaction time, depression score, difference in seated and standing diastolic blood pressure, and visual acuity in the left eye. The figure suggests that upper limits (red) of certain variables (e.g. chair stands time, age, body mass index, number of medications) are more negatively impactful than their lower limits, which are positively impactful. The opposite is the case for upper limits of grip strength, for example.

**Figure 3.**
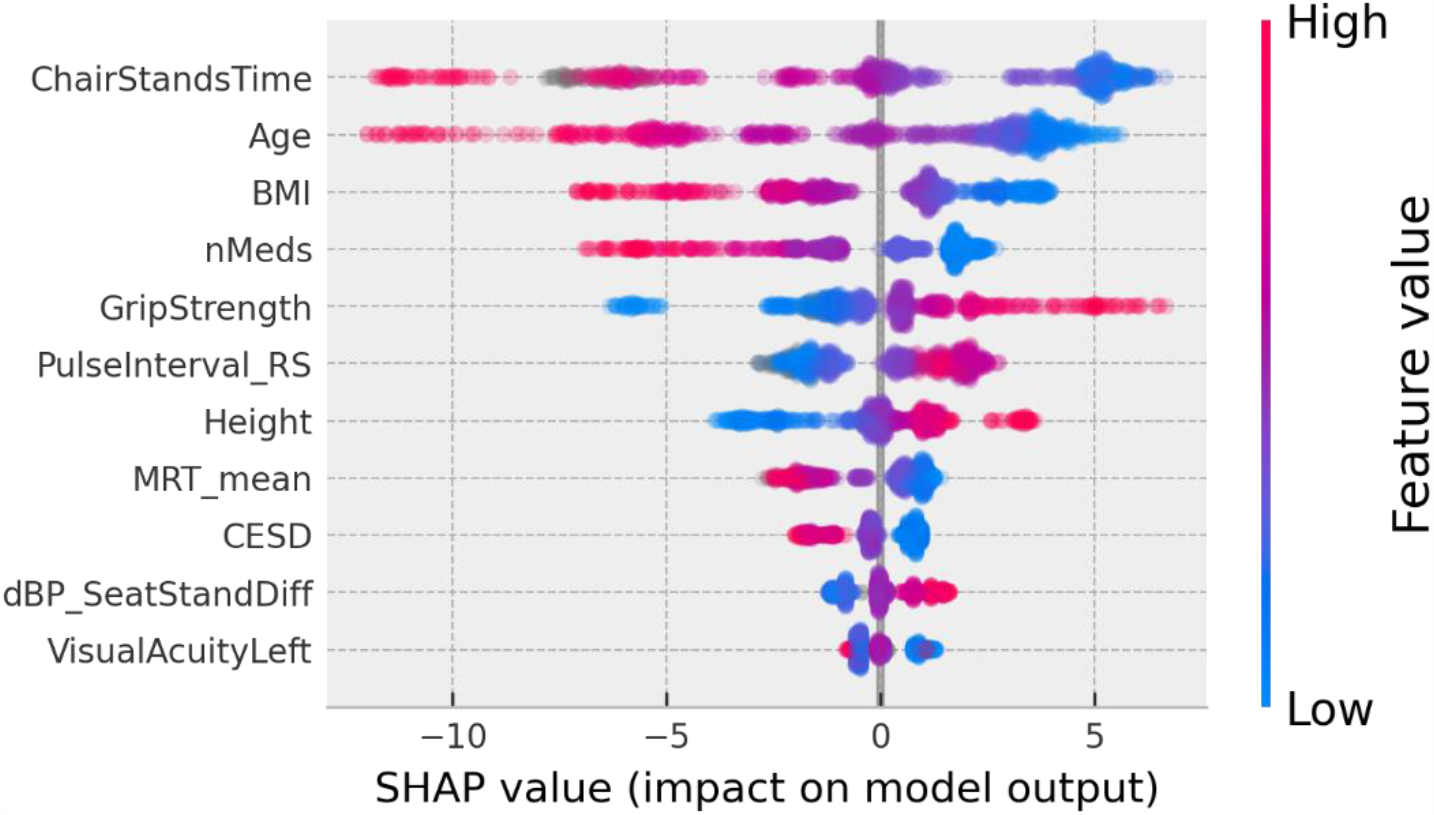
**SHAP summary plot for the final usual gait speed model. Features are ordered from top to bottom by decreasing mean absolute SHAP value. For each feature, each point represents a single sample in the test data. A sample’s x-coordinate displays the SHAP value for that sample with respect to a given feature. The colour of a sample indicates the value of the feature, with red being high, blue low, and grey missing.**

Scatter plots of SHAP value vs. feature can be seen for all features in Figure 4. SHAP values (left y-axis) vs. input feature value (x-axis) with underlaid histogram (right y-axis shows histogram counts) are shown for each feature in the UGS model. Features are arranged top-to-bottom and left-to-right in order of decreasing mean absolute SHAP value. At the zero point on the left y-axis (SHAP value = 0), the corresponding x-coordinate values for that feature are associated with having no impact on the model (i.e. they are associated with the mean model output). The vertical spread observed in the SHAP values vs. input feature plots indicates the presence of interaction effects. Although not chosen for the model the data points are coloured by sex.

**Figure 4.**
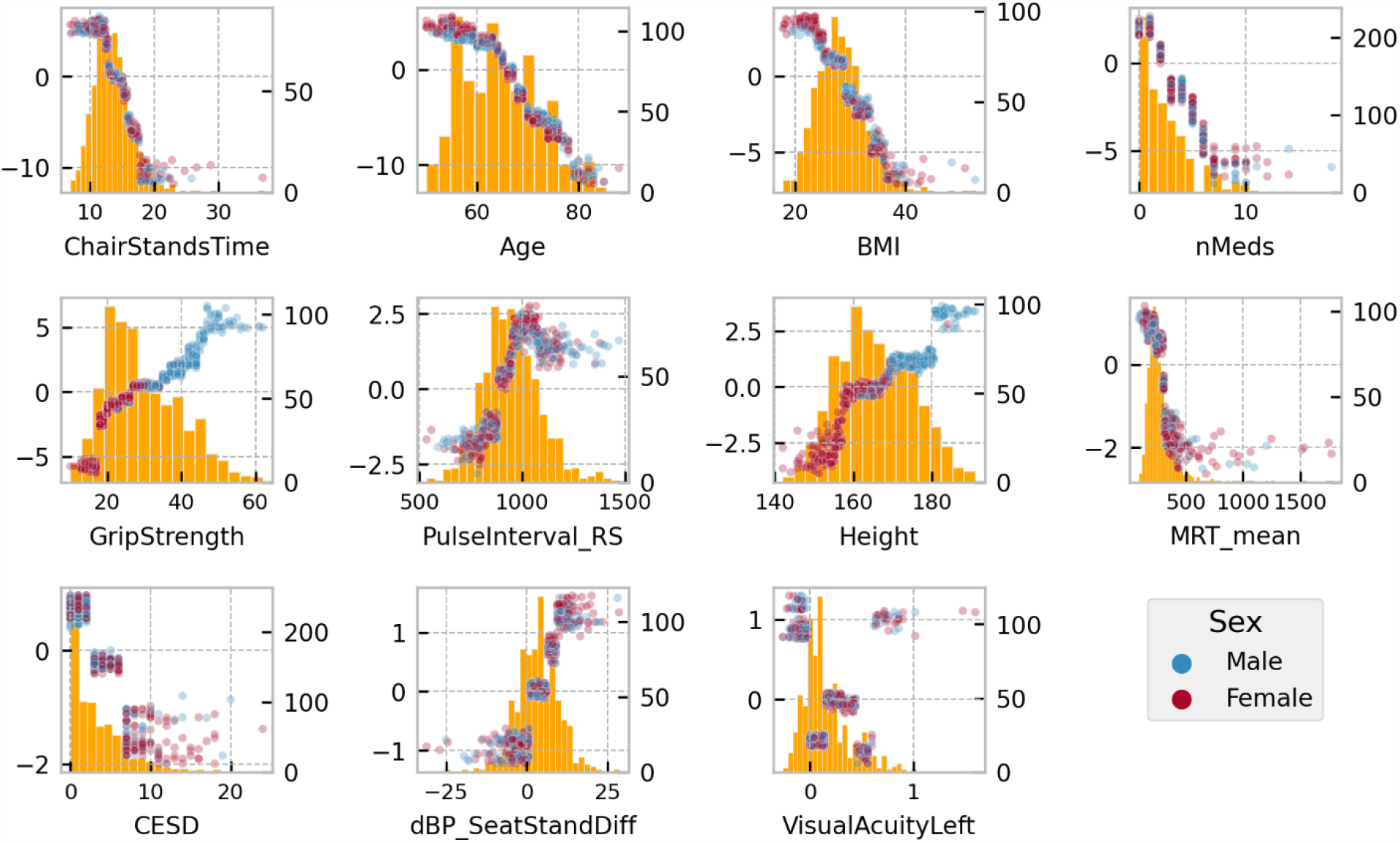
**SHAP values (left y-axis) vs. input feature value (x- axis) with underlaid histogram (right y-axis showing histogram counts) for each feature in the usual gait speed model. Features are arranged top to bottom and left to right in order of decreasing mean absolute SHAP value. Points are coloured be sex: male is blue and female is red.**

#### 3.2.1 Usual Gait Speed Model: Interactions

To further investigate the interaction effects suggested by vertical spreading in Figure 4, a plot of features ordered by decreasing mean absolute SHAP interaction value was produced (Figure 5); in it, features are ranked from left to right in order of decreasing mean absolute SHAP interaction values (blue line). Also shown in red are the mean maximum absolute SHAP interaction values, which can highlight the effects of outliers.

**Figure 5.**
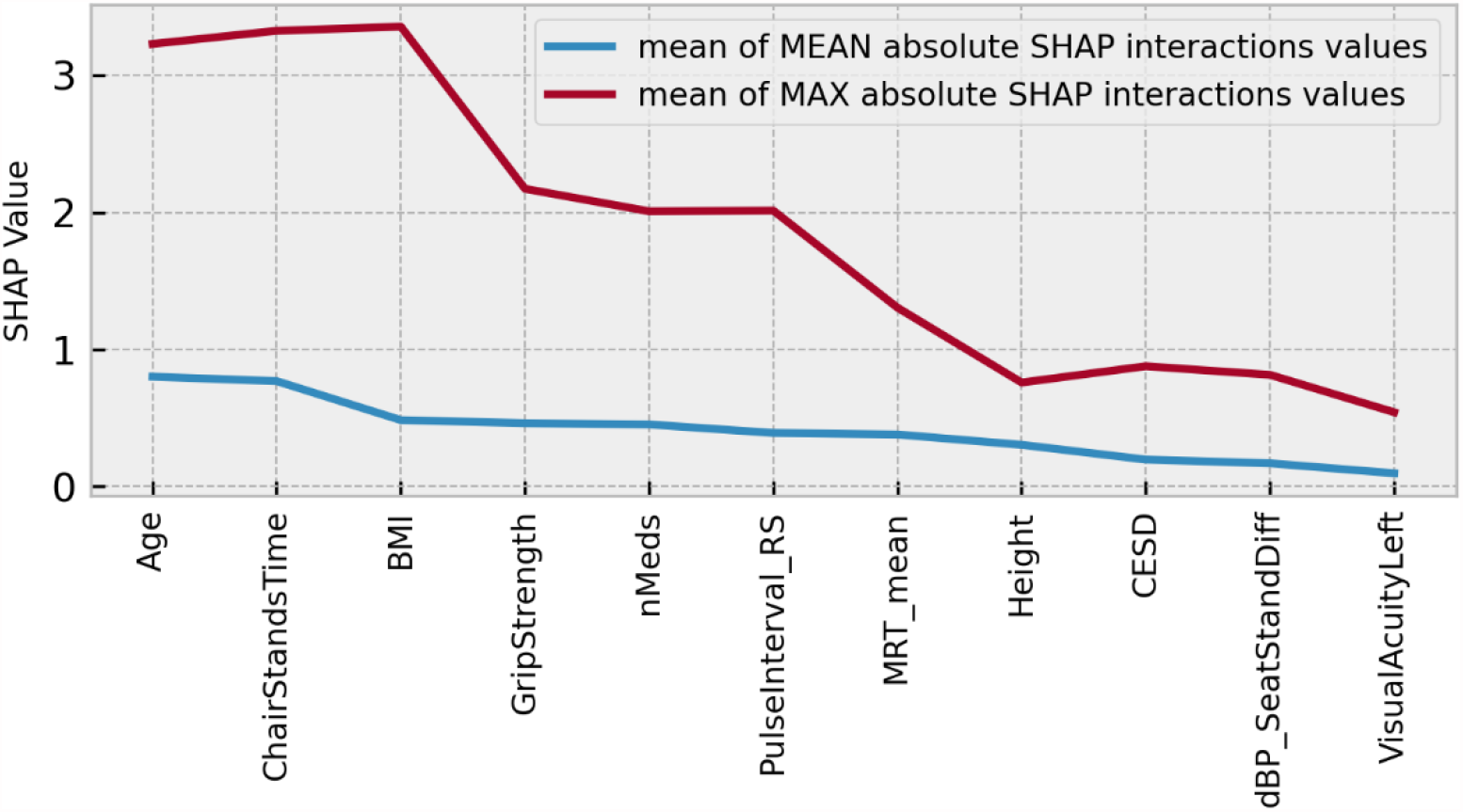
**Features ranked from left to right in order of decreasing mean absolute SHAP interaction values (blue line). Also shown in red are the mean maximum absolute SHAP interaction values, which can highlight the effects of outliers. Model: usual gait speed.**

The scatter plots of the top four interaction effects in the model (i.e. age, chair stands time, body mass index, and grip strength) are shown in Appendix 1. In the scatter plots, the points are coloured according to the value of the main interaction feature. The interactions are computed for the features in whatever numerical form they exist in, but for ease of visualization, continuous features are coloured according to what quartile a particular samples value falls in; blue indicates the value is in the lowest quartile and red the highest quartile. In each figure, the subplots are ordered from top-left to bottom-right by decreasing mean absolute SHAP interaction value.

### 3.3 Maximum Gait Speed

The peak 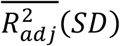 achieved for the MGS model was 0.45 (0.04), with training and test scores of 0.54 and 0.46, respectively. The expected model output was 170.9 cm/s. Features chosen for the model, in order of selection were: age, grip strength, chair stands time, body mass index, education, mean motor reaction time in the choice reaction time test, number of medications, height, the standard deviation of the mean reaction time in the sustained attention to response task, resting state heart rate, fear of falling, MOCA errors, orthostatic intolerance during active stand, smoking status, total power of the heart rate during paced breathing, the root mean square of successive differences between heartbeats during paced breathing, and best visual acuity. Figure 6 shows the visualization of the feature selection process for this model.

**Figure 6.**
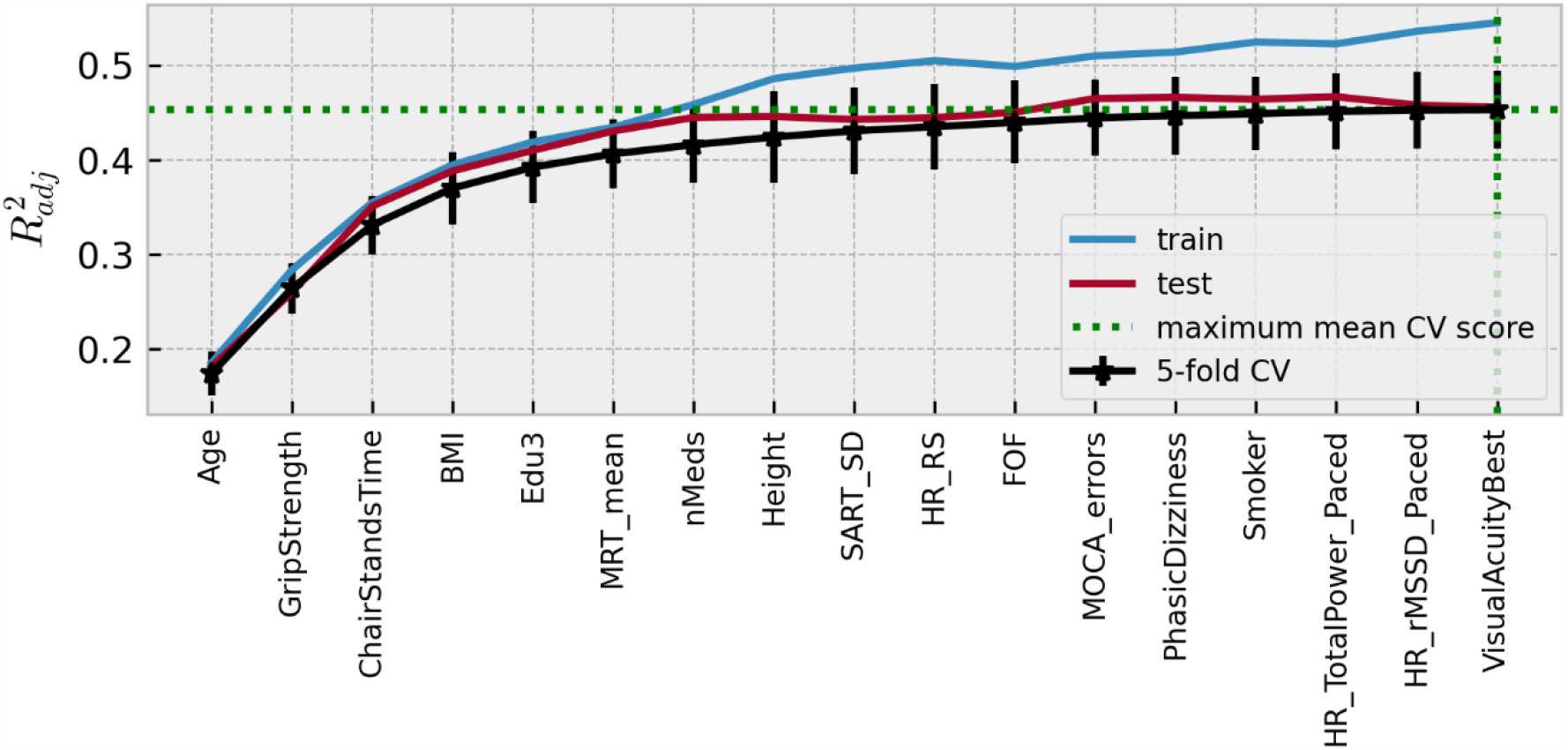
**Visualization of the feature selection process for the maximum gait speed model. From left to right on the x-axis, the features are in order of addition to the model. The y-axis shows the dimensionless** 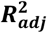 **metric**.**Mean 5-fold cross-validation scores with error bars showing ± SD are shown in black, train scores in blue, and test scores in red. The feature at which the peak score was achieved is highlighted by the green dotted line.**

In the SHAP summary plot for the MGS model shown in Figure 7, the feature importance ranked in order of decreasing mean absolute SHAP values was: age, chair stands time, grip strength, body mass index, height, number of medications, mean motor reaction time in the choice reaction time test, orthostatic intolerance during active stand, education, the standard deviation of the mean reaction time in the sustained attention to response task, fear of falling, MOCA errors, smoking, mean heart rate pre-active stand, the root mean square of successive differences between heartbeats during paced breathing, visual acuity, and total power of the heart rate during paced breathing.

**Figure 7.**
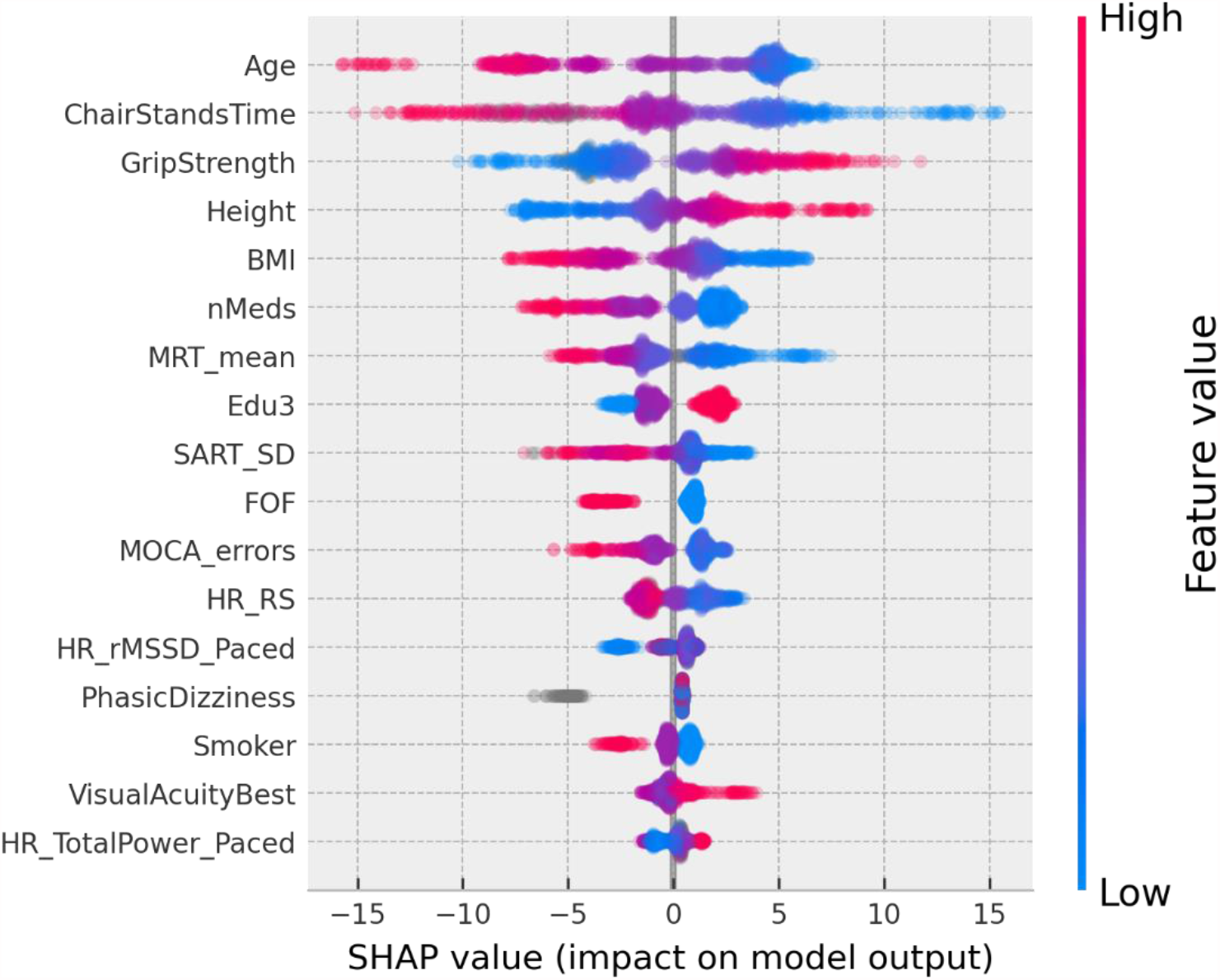
**SHAP summary plot for the maximum gait speed model. Features are ordered from top to bottom by decreasing mean absolute SHAP value. For each feature, each point represents a single sample in the test data. A sample’s x-coordinate displays the SHAP value for that sample with respect to the given feature. The colour of a sample indicates the value of the feature, with red being high, blue low, and grey missing.**

Figure 8 shows the SHAP values versus input feature values with underlaid histogram for each feature in the MGS model. Figure 9 shows a plot of features ordered by decreasing mean absolute SHAP interaction value, and Appendix 2 contains the scatter plots of the top four interaction effects in the model.

**Figure 8.**
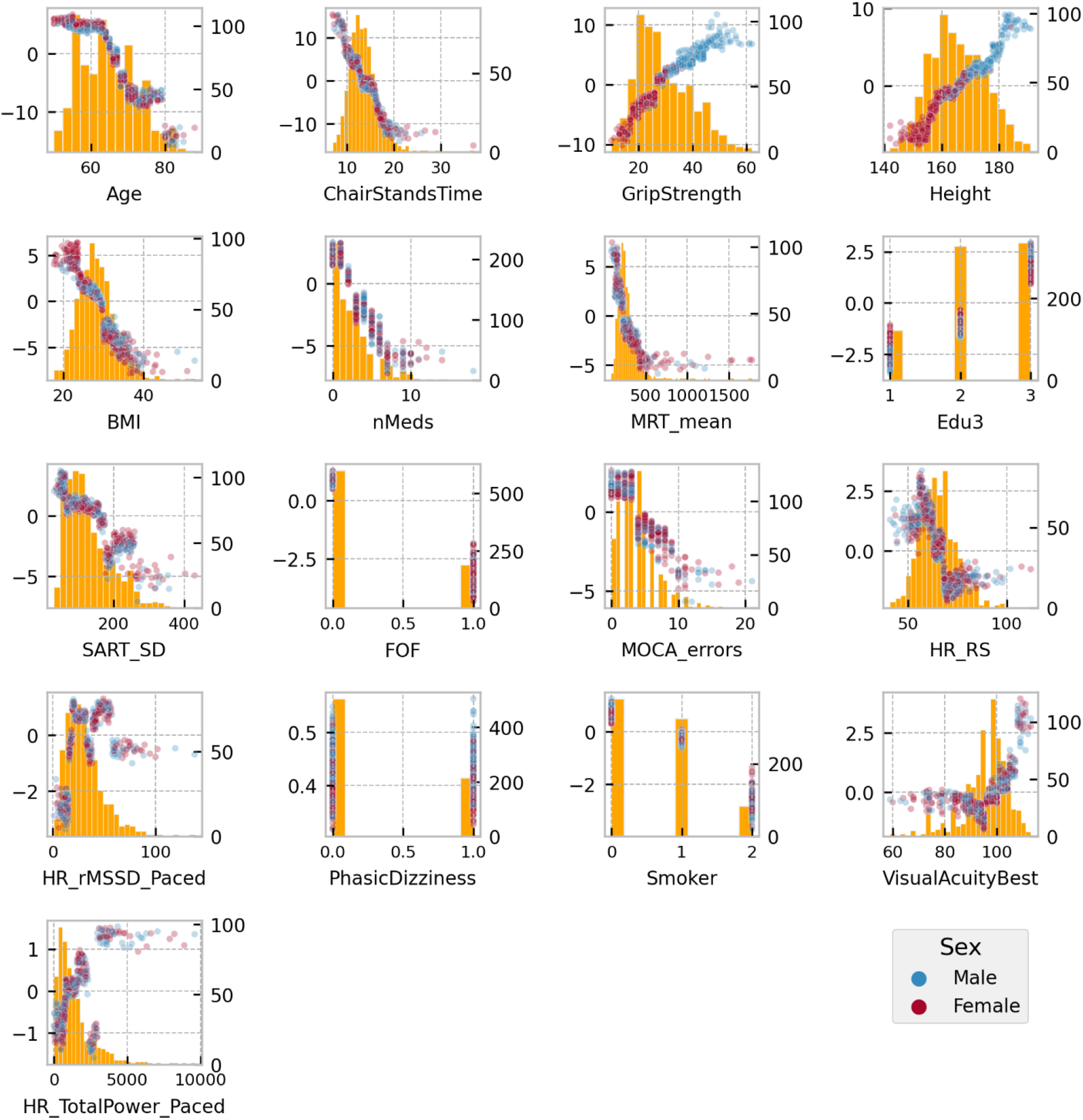
**SHAP values (left y-axis) vs input feature value (x- axis) with underlaid histogram (right y-axis shows histogram counts) for each feature in the maximum gait speed model. Features are arranged top to bottom and left to right in order of decreasing mean absolute SHAP value. Points are coloured be sex: male is blue and female is red.**

**Figure 9.**
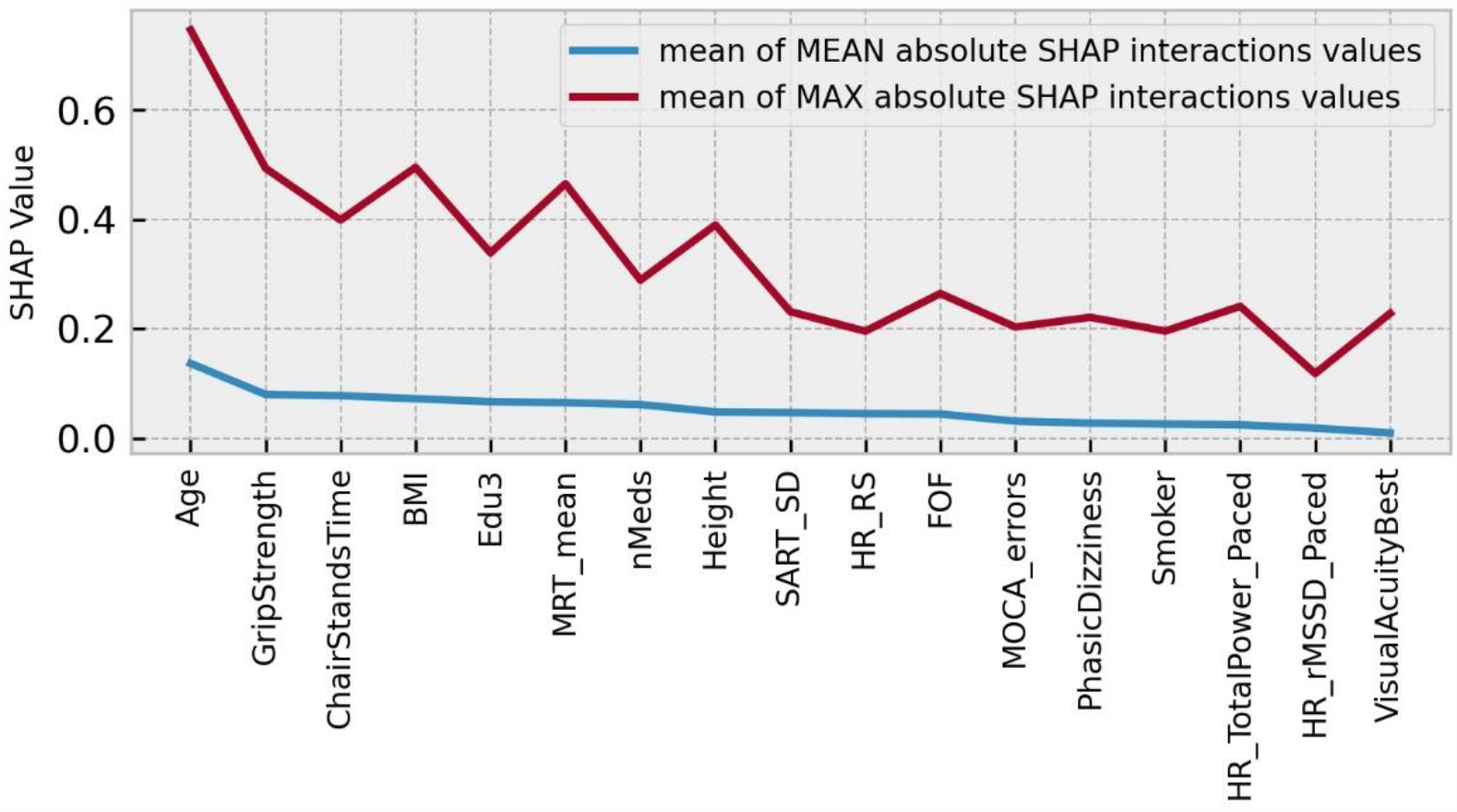
**Features ranked from left to right in order of decreasing mean absolute SHAP interaction values (blue line). Also shown in red are the mean max absolute SHAP interaction values which can highlight the effects of outliers. Model: maximum gait speed.**

### 3.4 Gait Speed Reserve

The peak 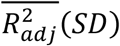 achieved for the GSR model was 0.19 (0.02), with training and test scores of 0.22 and 0.21, respectively. The model expected output was 34.2 cm/s. Figure 10 shows the visualization of the feature selection process. In order of selection, the features chosen were: mean motor reaction time in the choice reaction time test, grip strength, education, chair stands time, MOCA errors, accuracy proportion in the sound induced flash illusion (two beeps and one flash with stimulus-onset asynchrony of +150 ms), fear of falling, height, age, sex (0 = male; 1 = female), orthostatic intolerance in the active stand test, MMSE errors, and number of cardiovascular conditions.

**Figure 10.**
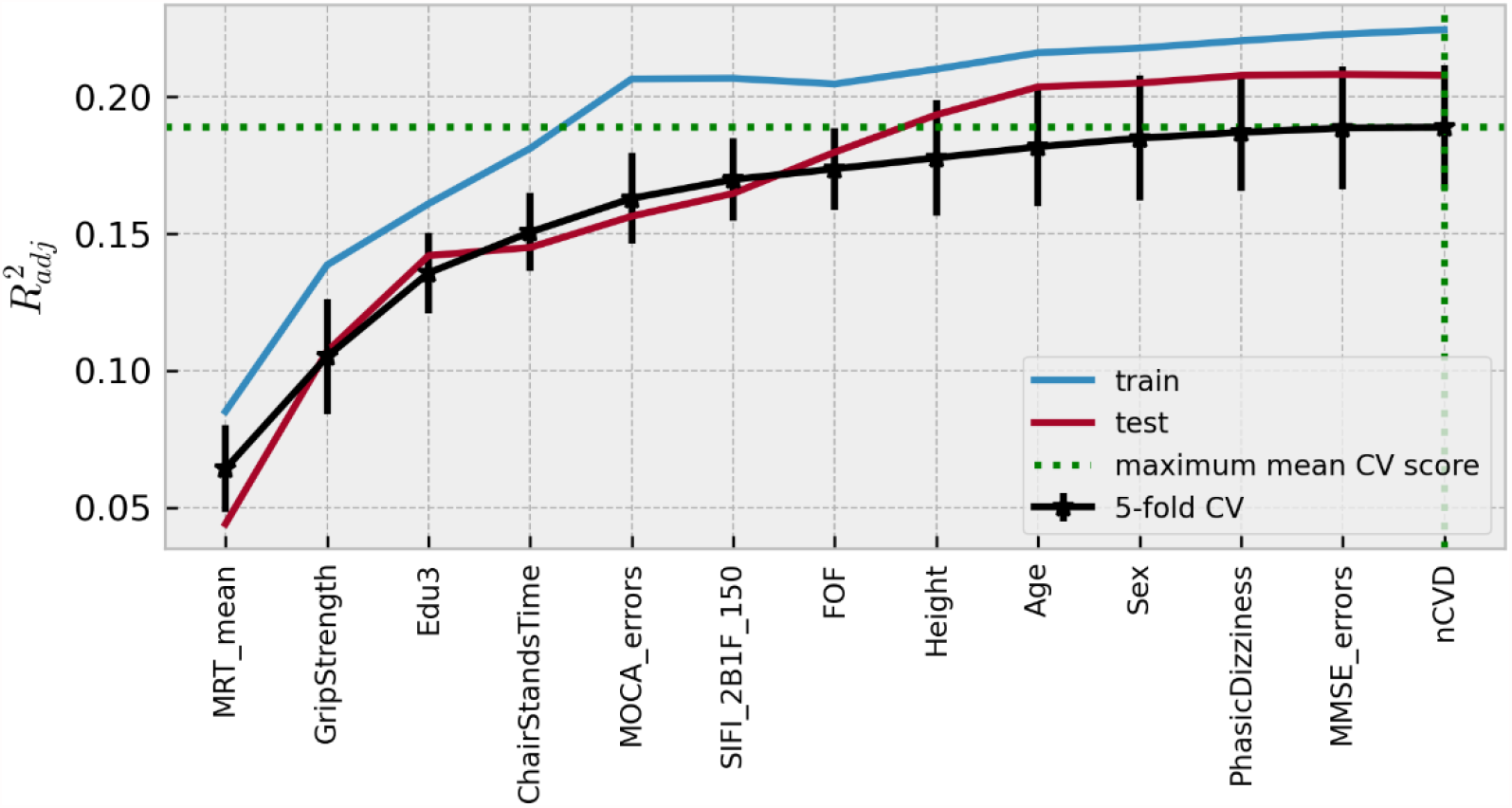
**Visualization of feature selection process for gait speed reserve. From left to right on the x-axis, the features are in order of addition to the model. The y-axis shows the dimensionless** 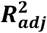 **metric. Mean 5-fold cross-validation scores with error bars showing ± SD are shown in black, train scores in blue, and test scores in red. The feature at which the peak score was achieved is highlighted by the green dotted lines.**

In the SHAP summary plot for the GSR model shown in Figure 11, the feature importance ranked in order of decreasing mean absolute SHAP values was: education, grip strength, mean MRT, MOCA errors, age, chair stands time, height, sex, accuracy proportion in the sound induced flash illusion, fear of falling, orthostatic intolerance, MMSE errors, and number of cardiovascular conditions.

**Figure 11.**
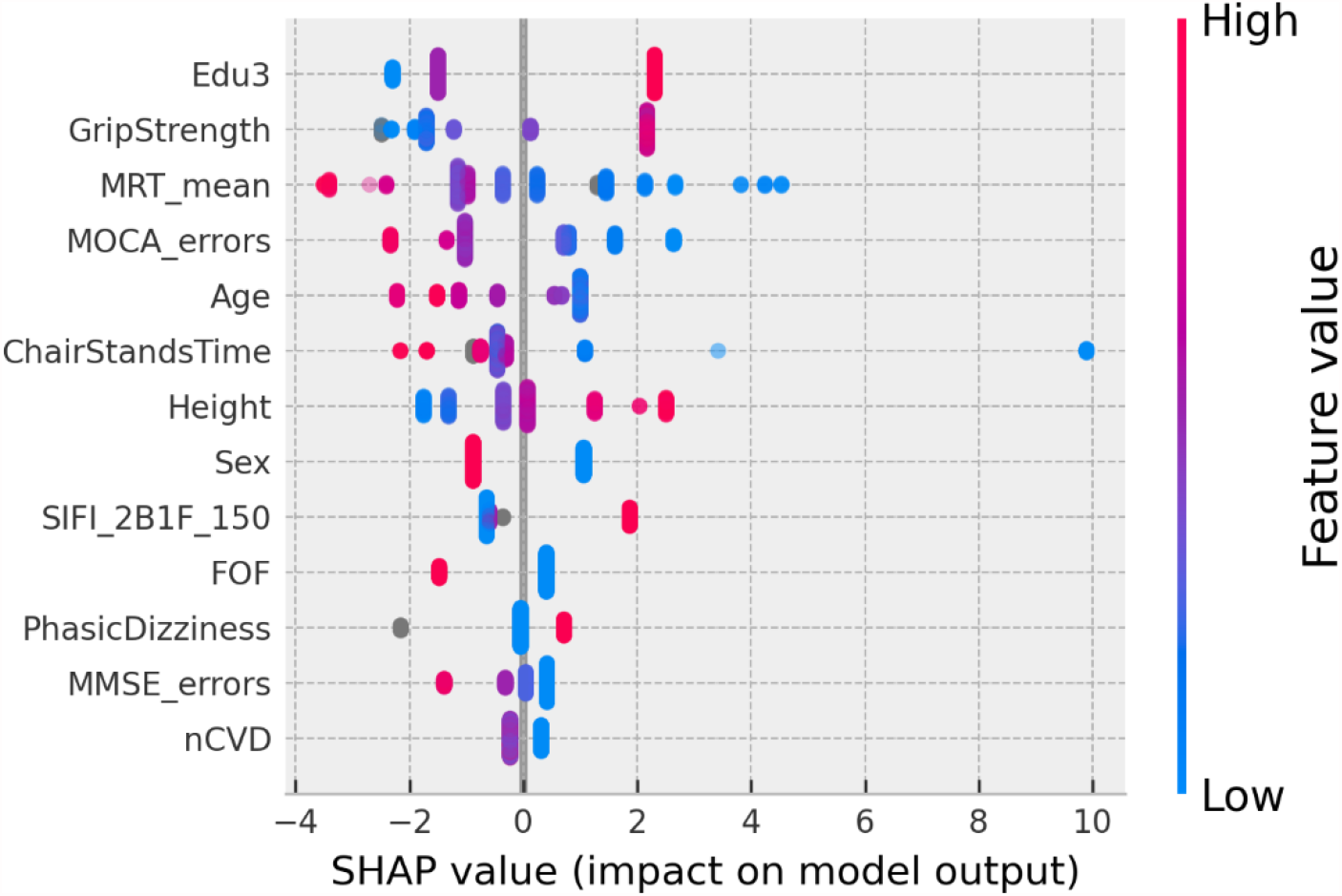
**SHAP summary plot for the final gait speed reserve model. Features are ordered from top to bottom by decreasing mean absolute SHAP value. For each feature, each point represents a single sample in the test data. A sample’s x-coordinate displays the SHAP value for that sample with respect to the given feature. The colour of a sample indicates the value of the feature, with red being high, blue low, and grey missing.**

Figure 12 shows the SHAP values versus input feature values with underlaid histogram for each feature in the GSR model. The absence of vertical spread in the SHAP vs. feature scatter plots is due to the maximum leaf nodes hyperparameter being set equal to two for the histogram gradient boosting model. This results in there being no interaction terms since the predictions made by each tree only considered features independently (i.e. a maximum leaf node limit of two means that for a given tree only a single split is made along a single feature).

**Figure 12.**
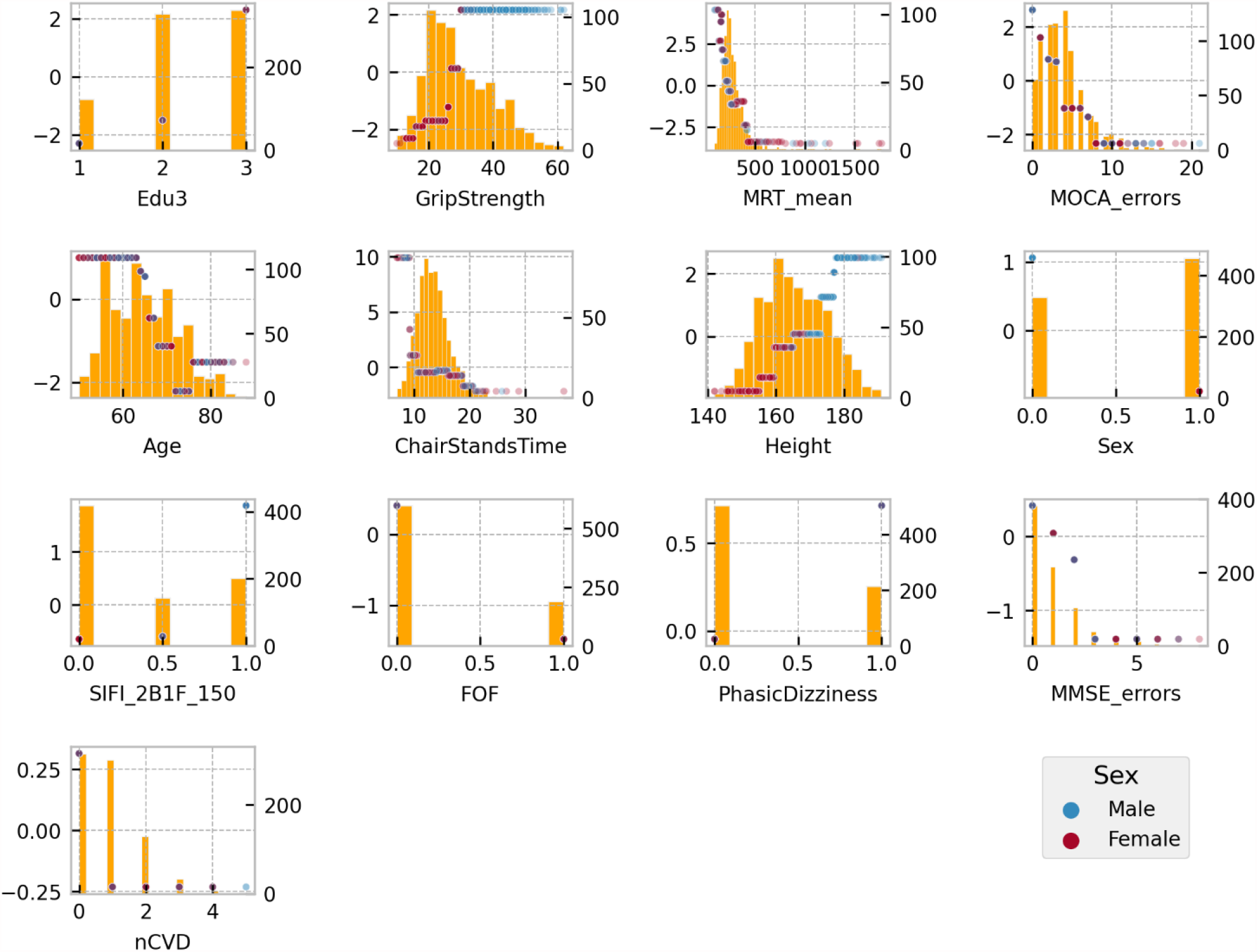
**SHAP values (left y-axis) vs input feature value (x- axis) with underlayed histogram (right y-axis shows histogram counts) for each feature in the gait speed reserve model. Points are coloured be sex: male is blue and female is red.**

The group mean differences in SHAP values along with 95% confidence intervals can be seen in Figure 13 for (A) sex, (B) third level education vs. all others, and (C) first/no education vs. all others. For sex, the grip strength feature produced a larger difference in means than sex itself with grip strength having a less positive impact for women. Height, mean MRT, fear of falling, and SIFI accuracy, were all significant and all exhibited a negative mean impact difference. On the other hand, for education there was a positive group mean difference for women in comparison to men. When comparing third/higher educational attainment to the rest, education itself seemed to make the only significant difference. However, when comparing primary/no educational attainment to secondary and tertiary educational attainment in Figure 13 (C) we observed several other significant differences other than education: MOCA errors, age, mean MRT, MMSE errors, illusion accuracy, orthostatic intolerance, fear of falling, and number of cardiovascular diseases.

**Figure 13.**
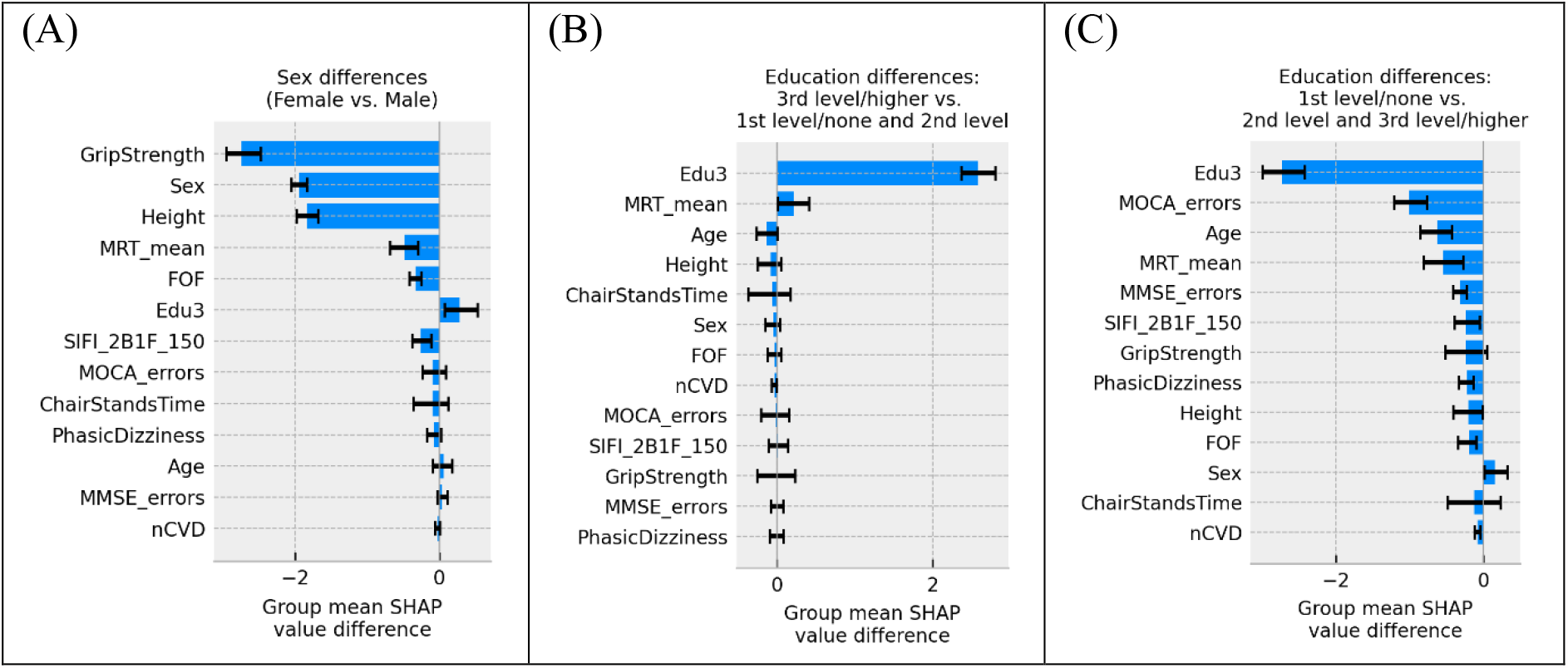
**Plots showing the group mean differences in SHAP values between subgroups with 95% confidence intervals. (A) shows the differences in sex, (B) shows the differences between participants with third/higher level of educational attainment and all others, and (C) shows the differences between participants with first/no level of educational attainment and all others.**

### 3.5 Summary of results

1 summarizes the scores and features selected for each models and Table 2 highlights the features that are unique to each model and those that are common to all three models.

**Table 1.**
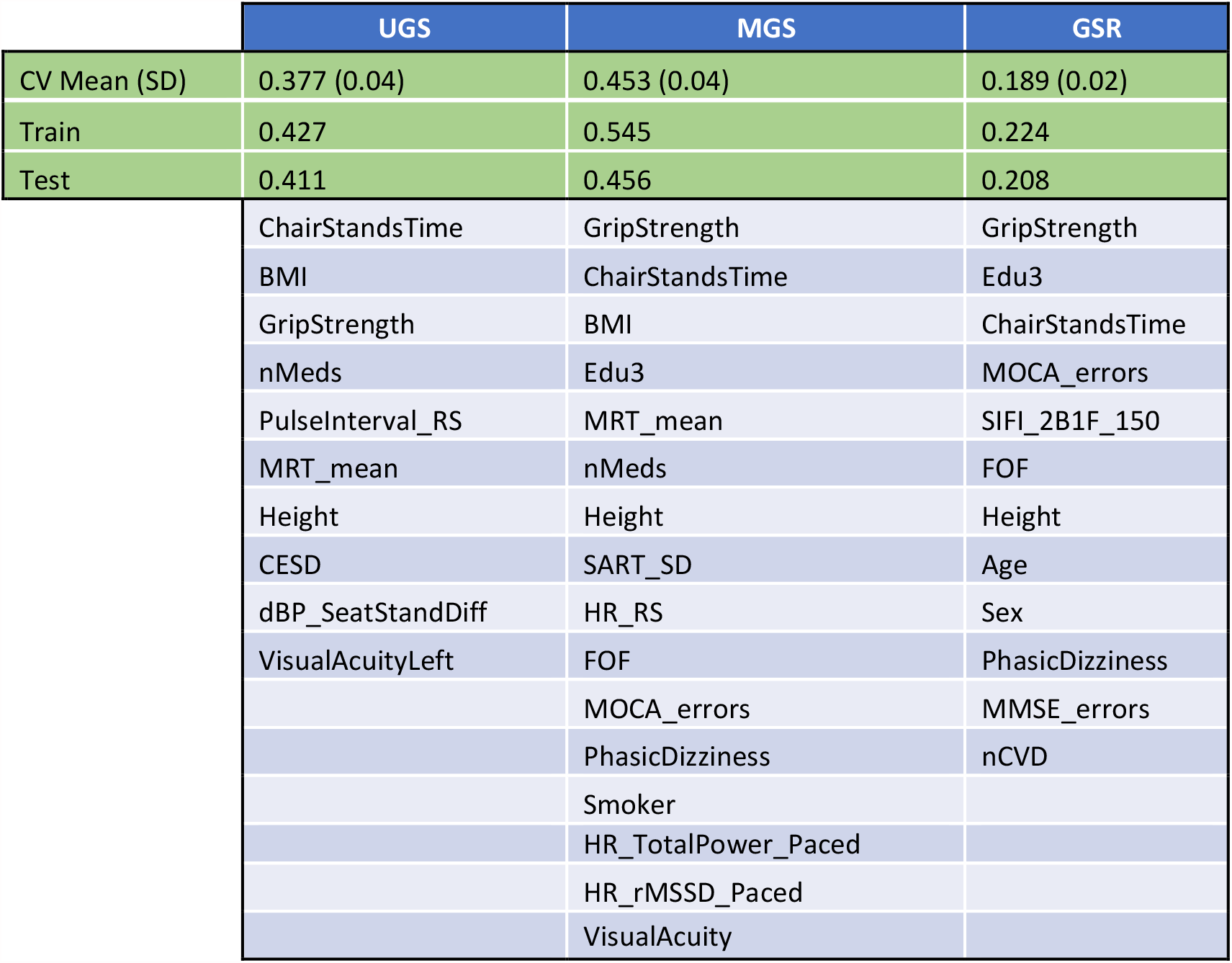
Summary of scores and features selected for the UGS, MGS, and GSR models.

|  | UGS | MGS | GSR |
| --- | --- | --- | --- |
| CV Mean (SD) | 0.377 (0.04) | 0.453 (0.04) | 0.189 (0.02) |
| Train | 0.427 | 0.545 | 0.224 |
| Test | 0.411 | 0.456 | 0.208 |
|  | ChairStandsTime | GripStrength | GripStrength |
|  | BMI | ChairStandsTime | Edu3 |
|  | GripStrength | BMI | ChairStandsTime |
|  | nMeds | Edu3 | MOCA_errors |
|  | PulseInterval_RS | MRT_mean | SIFI_2B1F_150 |
|  | MRT_mean | nMeds | FOF |
|  | Height | Height | Height |
|  | CESD | SART_SD | Age |
|  | dBP_SeatStandDiff | HR_RS | Sex |
|  | VisualAcuityLeft | FOF | PhasicDizziness |
|  |  | MOCA_errors | MMSE_errors |
|  |  | PhasicDizziness | nCVD |
|  |  | Smoker |  |
|  |  | HR_TotalPower_Paced |  |
|  |  | HR_rMSSD_Paced |  |
|  |  | VisualAcuity |  |

**Table 2.**
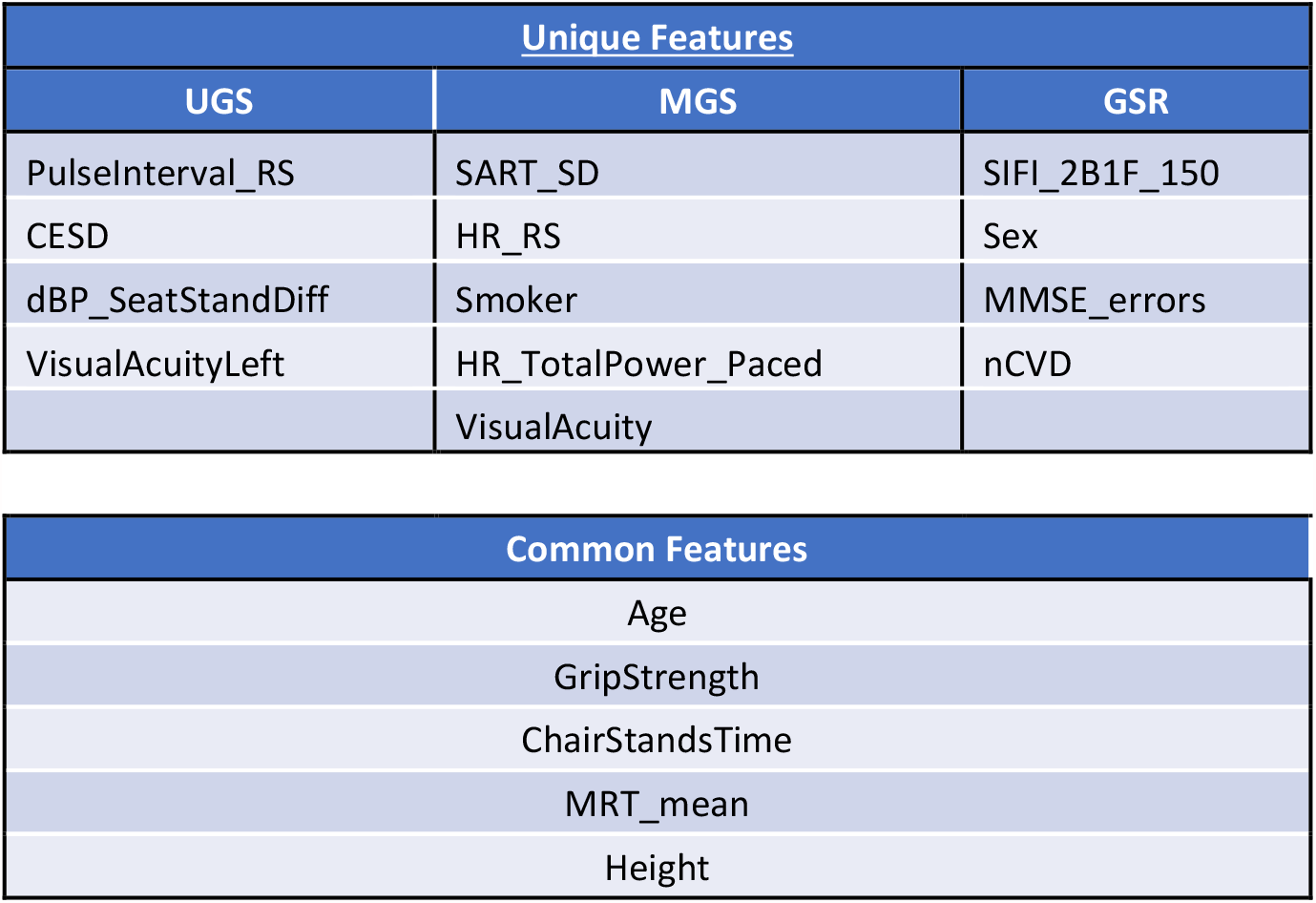
unique and common features across the UGS, MGS, and GSR models.

## 4 Discussion

### 4.1 Overall summary of findings

In the present study, using data from Wave 3 of TILDA, we used a gradient boosted trees-based stepwise feature selection pipeline for the discovery of clinically relevant predictors of GSR, UGS, and MGS using a shortlist of 88 features across 5 domains. The features selected for the respective models explained MGS and UGS to a greater extent than GSR. As shown in Table 2 there were common features, but also some features unique to each of the three models

### 4.2 Model Prediction

Based on model 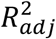 values, GSR (19%) was less predictable than MGS (45%) and UGS (38%). While we are not aware of previous published data for comparison with our GSR prediction, a previous study by Bohannon reported linear regression-based *R*^2^ values of 13% for UGS and 41% for MGS (44). Our results are comparably superior to those, especially given the fact that our *R*^2^ unit is adjusted and agree in that the MGS model yielded a larger prediction score than that of UGS.

### 4.3 Common Features

Across the three models, there were five common selected features: age, grip strength, chair stands time, mean motor reaction time in the choice reaction time test, and height. The top four features with the most impactful interactions (by mean absolute SHAP interaction value) were the same for the UGS and MGS models: age, chair stands time, grip strength, and body mass index.

Our results agree with Bohannon’s previous findings that UGS and MGS decline with increasing age (44). Other authors have also shown this for UGS (45-47). As per SHAP value vs. feature plots, increasing age was negatively associated with UGS, MGS, and GSR at ≥68, ≥68, and ≥66 years, respectively. Height is also unsurprising as a common predictor; indeed, taller people have longer legs and can achieve longer strides and higher velocity in any gait modality. Consequently, gait speed is often normalized by height (44, 48, 49).

It is also clinically plausible that higher grip strength (as a marker of upper limb strength) and shorter chair stands time (more representative of lower limb strength) were common determinants of all three performance metrics. Indeed, sarcopenia (low muscle mass and/or strength), of which both grip strength and the five chair stands test are indicative measures (50), has been associated with reduced gait speed and poor functional outcomes in older people (51-53). In our models, slower chair stands time was associated with a decline in UGS, MGS, and GSR once time increased beyond 14.2 s, 13 s, and 10.6 s, respectively; whilst increases in UGS, MGS, and GSR began at values of 13.4 s, 13 s, and 10.6 s, respectively. Grip strength of ≤26 kg was associated with slower UGS, MGS and GSR while grip strengths of ≥35 kg, 27 kg, and 27 kg, respectively, were associated with faster performance. These values for grip strength, while interesting from an absolute point of view, have a reduced clinical significance given the large differences in grip strength between men and women. Except for height, the other features relationships to the model output appear quite homogeneous with respect to sex.

Higher mean motor reaction time in the choice reaction time test was associated with lower speed in all three models. In previous research shorter CRT has been associated with faster gait speed after adjusting for potential confounders, and suggests that in older adults engaging more frequently in cognitively stimulating activities may improve neuromotor performance and mobility (55). In addition, our results resonate with previous TILDA work utilizing traditional linear statistics showing that participants in the slower MRT group (<250 ms) at Wave 1 seemed to have faster mobility decline as assessed by the timed up and go at Wave 3, four years later (29). Interestingly, in the latter study, the MRT cutoff was set arbitrarily, but in the present study the negative/positive impact thresholds for UGS, MGS and GSR were 299 ms, 231 ms, and 229 ms, respectively. Interestingly, the less physically demanding UGS model was only negatively influenced above a relatively slower MRT threshold.

The counter-intuitive results of higher grip strength, quicker chair stands time, and quicker MRT being associated with an increase in UGS when compared to MGS may be revealing of the underlying determining mechanisms of both acts; MGS may be a more physically determined act than UGS and easier to improve on than UGS.

Common between UGS and MGS models were BMI and number of medications, in the clinically expected directions, i.e. obesity and number of medications had a negative impact on gait speed. As regards obesity, research has suggested that obese adults may select their walking speed to minimize pendular energy transduction, energy cost, and perceived exertion during walking (56). In our UGS and MGS models, a BMI ≥29 kg/m^2^ had negative impact association. Hypothetically, it is possible that in TILDA, obese individuals equally reduced their UGS and MGS, which could possibly explain why BMI was not a feature in the GSR model. As regards number of medications, a similar mechanism could apply. In any case, our findings are in keeping with previous research showing that drug interactions may increase the likelihood of gait speed decline among older adults (57). In our UGS and MGS models, more than two medications had a negative impact association. The is below the usual polypharmacy definition of 5+ medication and the negative impact association with medications could be related to the underlying health condition rather than due to the medications themselves. Of note, visual acuity featured in both UGS (left) and MGS (best), but not in GSR, which could have a similar underlying reason (i.e. both UGS and MGS equally limited).

There were no features exclusively shared by UGS and GSR, but there were four features in the intersection of MGS and GSR: education, MOCA errors, fear of falling, and orthostatic intolerance. As regards the former two, tertiary education was associated with increased gait speed, and primary and secondary levels with a decrease. Greater than three MOCA errors negatively impacted both models. Interestingly, better MOCA performance is associated with higher education (58) and places greater emphasis on frontal executive function and attention tasks than the MMSE (59). Planning for the MGS task may require greater attention and executive function than performing the UGS task (60), and this may explain MOCA being related to GSR and MGS. Two or more MMSE errors were associated with GSR decrease.

Analogously, orthostatic intolerance and fear of falling may selectively limit the more demanding MGS task, but not the more comfortable UGS task. Orthostatic intolerance can be caused by orthostatic hypotension, which in some studies has been associated with reduced gait speed (61). In addition, orthostatic intolerance can be a feature of vestibular disorders such as benign paroxysmal positional vertigo (BPPV) (62), and research has suggested that the gait characteristics of BPPV can be attributed to an inadequate, cautious gait control (63), which may preferentially manifest in the MGS task. Fear of falling can also become stronger when facing the MGS task, compared to walking at UGS (64).

### 4.4 Unique features

Features exclusive to UGS were depression, diastolic blood pressure drop from sitting to standing, and resting state pulse interval. Higher levels of depressive symptoms have been associated with worse performance in specific quantitative gait variables in community-residing older adults, including lower velocity (65). In our model, CESD negatively impacted UGS when CESD>2 points.

Similarly, TILDA work showed that slower recovery of BP after standing (systolic and/or diastolic) was independently associated with poorer gait performance (61). On the other hand, a higher pulse interval indicates a higher heart rate variability and a more parasympathetic-driven autonomic cardiac control, which has been associated with healthier states (66) and mirrors the fact that for the UGS model, higher pulse intervals had positive influence. In our model, a baseline pulse interval of 799 ms or less had a negative impact on UGS (this is roughly 75.1 bpm: 60 seconds per minute / 0.799 seconds per beat).

Exclusive to the MGS model were the standard deviation of the mean reaction time in the sustained attention to response task, smoking, the mean heart rate pre-active stand, the total power of the heart rate during paced breathing, and the root mean square of successive differences between heartbeats during paced breathing. In a previous study, community-dwelling participants who displayed poorer sustained attention walked more slowly during both single and dual gait tasks (67). In our model, standard deviation of the mean SART reaction time <157.7 ms was associated with slower MGS. Interestingly, research has shown that in habitual smokers, smoking acutely reduces baseline levels of vagal-cardiac nerve activity and completely resets vagally mediated arterial baroreceptor-cardiac reflex responses (68), which could be in keeping with heart rate and heart rate variability features being selected in this model. A baseline heart rate of 67.9 bpm or more had a negative impact on MGS in our model. Comparing this to the pulse interval of 799 ms (equivalent to 75.1 bpm) associated with the beginning of negative impact association in the UGS model, we see that in terms of an increasing heart rate MGS begins to decline earlier than UGS.

Finally, features exclusive to GSR were accuracy proportion in the sound induced flash illusion (two beeps and one flash with stimulus-onset asynchrony of +150 ms), sex, MMSE errors, and number of cardiovascular diseases. Male sex was associated with increased GSR, potentially because men may comparatively accelerate more than women during the MGS task. Alternatively, this may also be because the variance explained by the GSR model was relatively low and the effect of sex might disappear when additional features are selected as in other models. One or more cardiovascular diseases was negatively associated with GSR, which is in keeping with the possibility that this type of disease may limit MGS more than UGS. As noted by a previous study (69), the difference between UGS and MGS is predominantly dictated by the latter. A notable exclusive associate of GSR was the proportion of accuracy in the sound induced flash illusion. This can be interpreted in the context that worse visual– somatosensory integration is associated with worse balance in older people (70), and that an increase in susceptibility to the sound-induced flash illusion during standing relative to sitting was present in fall-prone older adults (71).

### 4.5 Strengths of the Study

Strengths of the study include the use of a machine learning model that bins values for faster computation; offers native support for categorical features without the need for one-hot encoding (dummy variables); has native support for missing values not requiring removal of features/samples or imputation procedures; obviates the need to scale features as it is based on decision trees; allows for non-linear relationships; and makes no assumptions about underlying structure.

Another strength of the explainable machine learning methodology is that in the SHAP value verses feature plots, one can recognize the presence of what could be considered as ‘floor’ and ‘ceiling’ effects in the features. This highlights the importance of using non-linear models in this type of research, as even if the relationship observed within the ‘active’ region of the feature is indeed linear, a linear model cannot detect the plateau regions and would instead return a model coefficient that underestimates the effect size in the ‘active’ region. Clinical cut-offs and regions of interest for certain features are identifiable, as we have detailed above, making the models highly interpretable for clinicians.

### 4.6 Limitations of the Study

However, while these cut-offs and regions of interest may be able to inform the clinician it is possible that they may vary between populations. In terms of the analytical sample, this only included TILDA Wave 3 participants who underwent the health centre assessment where gait speed tests were conducted. Even though at Wave 1, TILDA was designed as a nationally representative cohort of people aged 50 or more years living in Ireland (16), our analytical sample at Wave 3 is not population-representative, and therefore our results are not necessarily generalizable to the Irish population. Indeed, TILDA work showed that participants attending the health assessment centre were generally fitter than those having a health assessment in their homes (72), which means that other features may have been selected in the models should frailer people have been included in the analytical sample.

Despite having many advantages, the machine learning methodology also has limitations. The features selected need to be considered in terms of the ‘package’ of features chosen for the final model, and not necessarily in hierarchical order of importance. It cannot be assumed that features not chosen for a model are not also predictive of the outcome variable. Even though measures were put in place to help reduce overfitting, in the absence an external validation sample, the risk of overfitting still exists. The confidence intervals of the effects and associations are also not known in this work however application of bootstrapping methods may be used in future work to address this.

Furthermore, the models are dependent on the predictors that were entered. Even though the ‘shortlist’ of predictors was quite comprehensive (i.e. 88 features across 5 domains), we may not have considered potentially relevant predictors that were either not measured or not shortlisted. In view of GSR being less predictable than UGS and MGS, it is possible that including additional features in the GSR model (perhaps personality/social/lifestyle factors) would improve the model prediction. Height-normalized gait speed could have been considered in the models, but this is not something that we wanted to consider a priori given the data-driven approach.

Another limitation touched on in the discussion is regarding the sex differences in grip strength and height. Height is not too much of an issue as it is non-modifiable and is a common choice for gait speed normalization; but the thresholds observed in grip strength with respect to positive or negative deviation from the mean in UGS, MGS, or GSR are heavily distorted by sex. A sex stratified investigation of grip strength in this context may be of clinical benefit given its modifiable nature and its high importance in all three models.

Finally, it must be made clear that despite the use of word ‘impact’ when explaining the relationship of input features to the output, all results are associations and causal relationships cannot be assumed.

### 4.7 Conclusions

The selected variables explained a greater proportion of variation in MGS and UGS than GSR. There were common features to all three models (i.e. age, grip strength, chair stands time, mean motor reaction time in the choice reaction time test, and height), but also some unique features to each of them. Overall, findings on all three models were clinically plausible and support a network physiology approach (73) to the understanding of predictors of performance-based tasks. Each model contains features from multiple physiological systems and thus support the hypothesis that GSR as well as UGS and MGS are multisystem phenomena. By employing an explainable machine learning model, our observations may help clinicians gain new insights into the possible determinants of physiological reserve in older adults. Of the features selected some are non-modifiable e.g. age, sex, height. Others however may be directly modifiable through changes in lifestyle, engaging in physical exercise, or cognitive stimulation (e.g. BMI, weight, smoking, education, chair stands time, grip strength, MOCA, motor response time, SART). For some variables it may be useful to focus on ensuring that a patient avoids reaching threshold values that are associated with a rapid decline in gait speed. Conversely, if engaging in rehabilitation those threshold values may be the targets so as to reach a more stable situation with respect to walking speed. Having explored the predictors of GSR and found multisystem associations further work will investigate whether GSR is a useful measure in predicting adverse health outcomes and if it can contribute to informing on overall physiological reserve.

## Data Availability

The datasets generated during and/or analysed during the current study are not publicly available due to data protection regulations but are accessible at TILDA on reasonable request. The procedures to gain access to TILDA data are specified at https://tilda.tcd.ie/data/accessing-data/.

https://tilda.tcd.ie/data/accessing-data/

## Funding

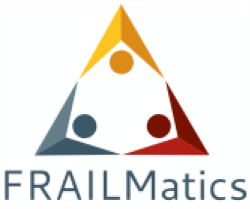

This research was funded by Science Foundation Ireland (SFI), grant number 18/FRL/6188. The Irish Longitudinal Study on Ageing (TILDA) is funded by Atlantic Philanthropies, the Irish Department of Health and Irish Life.

All calculations were performed on the Tinney cluster maintained by the Trinity Centre for High Performance Computing (Research IT) (https://www.tchpc.tcd.ie/node/1353). This cluster is funded form a Grant from Science Foundation Ireland under Grant number 18/FRL/6188.

