## Appendix 1 for "Comparison of gait speed reserve, usual gait speed, and maximum gait speed of adults aged 50+ in Ireland using explainable machine learning"

Scatter plots of the four top interaction effects on the **usual gait speed model** (i.e. age, chair stands time, body mass index, and grip strength). In the scatter plots, the points are coloured according the value of the main interaction feature. The interactions are computed for the features in whatever numerical form they exist in but for ease of visualization, continuous features are coloured according to what quartile a particular sample's value falls in; blue indicates the value is in the lowest quartile and red the highest quartile. In each figure, the subplots are ordered from top-left to bottom-right by decreasing mean absolute SHAP interaction value.

### Age Interactions

The SHAP values explaining the effect of interaction between age and the other features in the model are presented below. SHAP values for the interaction are on the y-axis, and feature values are on this x-axis; the colour of a point represents the age for that point, with blue being younger and red being older.

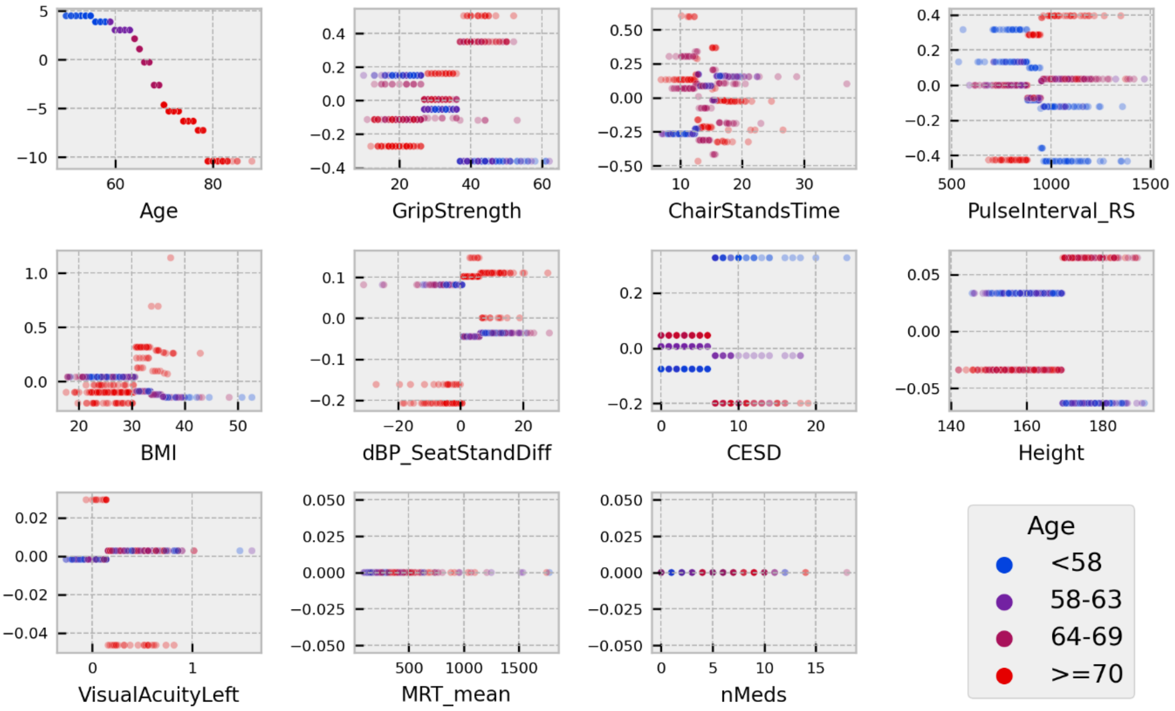


**Figure 1 - SHAP values for the interaction between age and all features in the usual speed model. The x-axes present the values of a feature in the units of that feature. The colour of a point indicates what quartile for age that sample falls in. The y-axis displays the SHAP value of the interaction.**

### Chair Stands Time Interactions

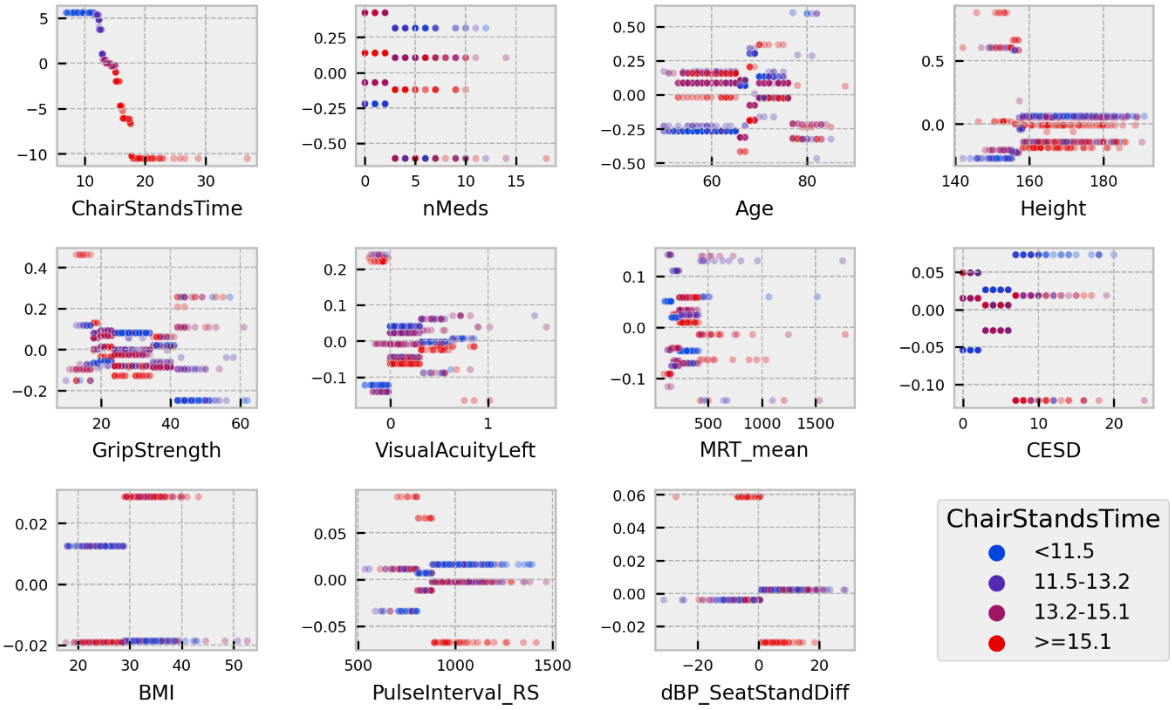


**Figure 2 - SHAP values for the interaction between chair stands time and all features in the usual speed model. The x-axes present the values of a feature in the units of that feature. The colour of a point indicates what quartile for chair stands time that sample falls in. The y-axis displays the SHAP value of the interaction.**

### BMI Interactions

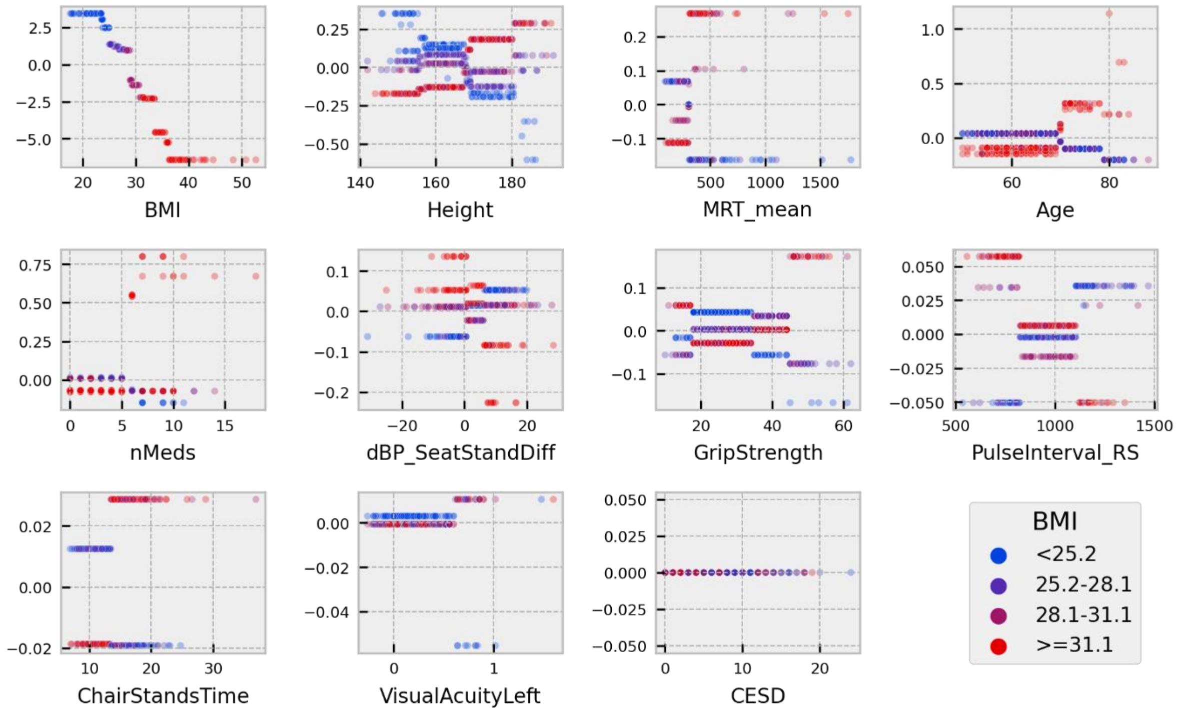


**Figure 3 - SHAP values for the interaction between BMI and all features in the usual speed model. The x-axes present the values of a feature in the units of that feature. The colour of a point indicates what quartile for BMI that sample falls in. The y-axis displays the SHAP value of the interaction.**

### Grip Strength Interactions

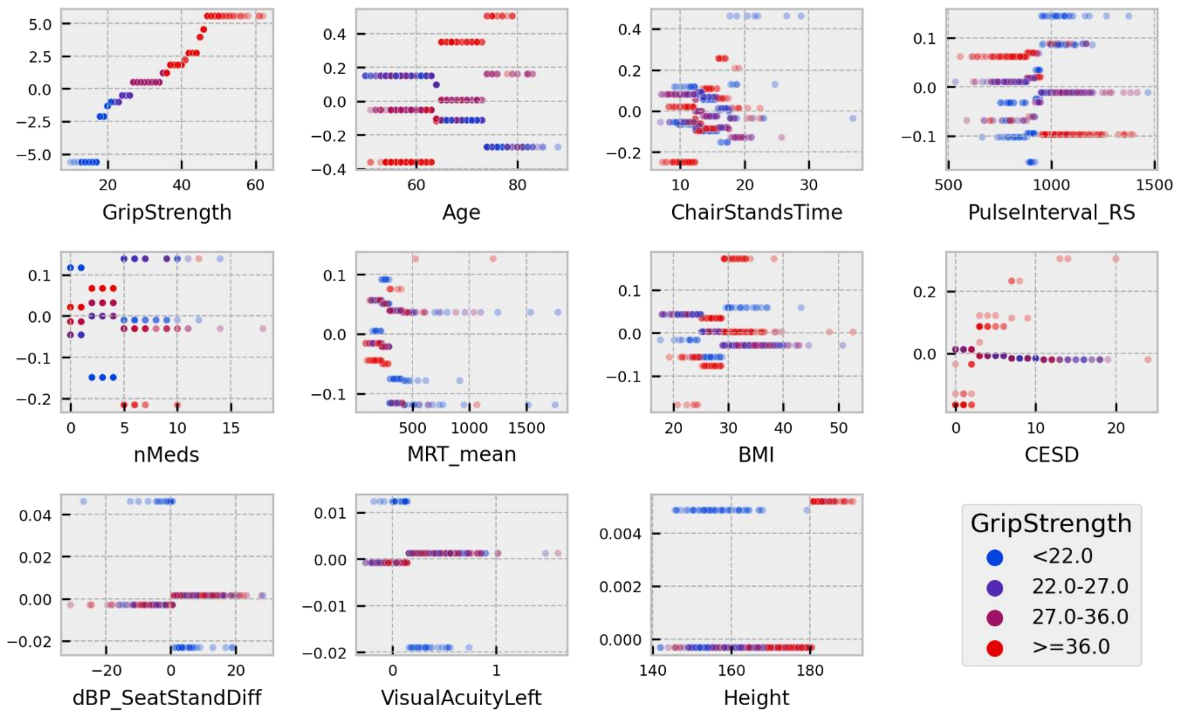


**Figure 4 - SHAP values for the interaction between grip strength and all features in the usual speed model. The x-axes present the values of a feature in the units of that feature. The colour of a point indicates what quartile for grip strength that sample falls in. The y-axis displays the SHAP value of the interaction.**
